# Effect of COVID-19 vaccination on mortality by COVID-19 and on mortality by other causes, the Netherlands, January 2021- January 2022

**DOI:** 10.1101/2022.07.21.22277831

**Authors:** Brechje de Gier, Liselotte van Asten, Tjarda M. Boere, Annika van Roon, Caren van Roekel, Joyce Pijpers, C.H. (Henri) van Werkhoven, Caroline van den Ende, Susan J.M. Hahné, Hester E. de Melker, Mirjam J. Knol, Susan van den Hof

## Abstract

**Background:** We aimed to estimate vaccine effectiveness (VE) against COVID-19 mortality, and to explore whether an increased risk of non-COVID-19 mortality exists in the weeks following a COVID-19 vaccine dose.

**Methods:** National registries of causes of death, COVID-19 vaccination, specialized health care and long-term care reimbursements were linked by a unique person identifier using data from 1 January 2021 to 31 January 2022. We used Cox regression with calendar time as underlying time scale to, firstly, estimate VE against COVID-19 mortality after primary and first booster vaccination, per month since vaccination and, secondly, estimate risk of non-COVID-19 mortality in the 5 or 8 weeks following a first, second or first booster dose, adjusting for birth year, sex, medical risk group and country of origin.

**Results:** VE against COVID-19 mortality was >90% for all age groups two months after completion of the primary series. VE gradually decreased thereafter, to around 80% at 7-8 months post-primary series for most groups, and around 60% for elderly receiving a high level of long-term care and for people aged 90+ years. Following a first booster dose, the VE increased to >85% in all groups. The risk of non-COVID-19 mortality was lower or similar in the 5 or 8 weeks following a first dose compared to no vaccination, as well as following a second dose compared to one dose and a booster compared to two doses, for all age and long-term care groups.

**Conclusion:** At the population level, COVID-19 vaccination greatly reduced the risk of COVID-19 mortality and no increased risk of death from other causes was observed.

## Background

COVID-19 vaccination programmes aim to reduce severe disease and death from COVID-19. Therefore, risk groups for severe outcomes of COVID-19 were prioritized for COVID-19 vaccination in many countries, including the Netherlands. Previous studies on vaccine effectiveness (VE) against death due to COVID-19 have shown high VE shortly after the primary series, with waning of VE over time since vaccination (1, 2, 3, 4, 5, 6).

Monitoring of vaccine safety has revealed that the majority of known side effects of COVID-19 vaccines are mild and self-limiting. Severe adverse events are very rare (7). Nonetheless, fatal consequences of severe adverse events after COVID-19 vaccination have been reported, such as thrombosis combined with thrombocytopenia after vaccination with viral vector vaccines (8, 9). Especially for people not belonging to risk groups for severe COVID-19, continuous monitoring of the risk-benefit ratio of vaccination is of utmost importance.

We aimed to assess associations between COVID-19 vaccination and mortality for the Dutch population during the study period from 1 January 2021 to 31 January 2022 in two ways. First, VE against death from COVID-19 was estimated. Second, risk of death from causes other than COVID-19 during the weeks immediately following a first, second or booster dose of vaccination was compared to that of the previous vaccination status (no vaccination, one or two doses, respectively).

## Methods

### Study population

Several national databases were linked within the secure remote access research platform provided by Statistics Netherlands using a unique personal identifier. All results are based on calculations by the authors using non-public microdata from Statistics Netherlands. All registered inhabitants of the Netherlands on 1 January 2021, born in or before 2009 (age ≥11 years), were included in the cohort. The population registry data (containing date of birth and death, sex, country of origin), using a unique personal identifier, were linked to the national COVID-19 vaccination registry CIMS (extraction date 3 May 2022), cause of death data (up to 31 January 2022), long-term care (LTC) reimbursement data (extraction date 2 February 2022), and outpatient hospital care claims registry (2016-2020 data) plus medication dispensation data (2020 data) to define medical risk.

### Vaccination status

With each administered COVID-19 vaccine in the Netherlands, the recipient is asked for consent for central registry of the vaccination in CIMS. Around 93% of persons receiving primary vaccination and 95% of persons receiving booster vaccination gave consent for registration. The date, product and series label (primary or booster dose) from CIMS were used to define start and end dates of vaccination statuses per person. As COVID-19 vaccination in the Netherlands commenced on January 6, 2021, each person started with the status ‘unvaccinated’ at 1 January 2021. Persons without registered vaccinations in CIMS retained the status ‘unvaccinated’. Vaccinees who did not give informed consent could not be distinguished from those who were not vaccinated.

For the VE analyses, vaccination status was defined as follows. The status “Primary series – partly” started on the day of the first administered dose. Fourteen days after the second dose of Comirnaty (Pfizer/BioNtech), Spikevax (Moderna) or Vaxzevria (AstraZeneca/ Oxford University), or 28 days after a single dose of Janssen vaccine, status “Primary series – completed” started, in line with the requirements for the COVID-19 vaccination certificate (10). Seven days after receipt of a booster dose, status “Boosted” commenced (11). For 0.5% of all people with vaccinations registered in CIMS, only a booster dose but no primary series was registered (presumably because no consent was given for registration of the primary series). In these cases, the first 6 days after receipt of the booster dose were marked as “Primary series – completed” (and as “Boosted” thereafter).

For the analyses regarding risk of death following vaccination, exposure statuses were pre-determined after consultation with experts from the National Pharmacovigilance Center. The eight weeks immediately following vaccination were defined as ‘risk periods’ and each week was analyzed separately. After a first dose of an mRNA vaccine, only the first five weeks (instead of 8 weeks) were analyzed as risk periods, because (generally) after 5 weeks, the second dose was administered. Unvaccinated person-time was used as reference for the analysis of risk of death after the first dose of COVID-19 vaccine. For the relative risk of death after a second dose, person-time at more than 14 days after a first dose was used as reference. For the relative risk of death after booster vaccination, person-time at least 3 months after a second dose was used as reference, because individuals were only eligible for booster vaccination 3 months or longer after a primary series. Persons who had received a 3-dose primary series due to an immunocompromising condition (N = 133,380) were excluded from the analysis of risk of death after booster vaccination, as high death rates in this especially vulnerable population might deflate hazard ratio estimates after the booster.

### Cause of death

Underlying cause of death, as reported by a physician on death registry forms and coded centrally per WHO guidelines by Statistics Netherlands, was available as ICD-10 codes. ICD-10 codes U07.1 and U07.2 were used to define COVID-19 mortality, which was the outcome for the VE analyses. All other codes were used to define non-COVID-19 mortality, which was the outcome for the analyses regarding risk of death following vaccination.

### Year of birth, sex, country of origin

Age was estimated as 2021 minus birth year. If a person was born abroad, their ‘country of origin’ was the country of birth. If a person was born in the Netherlands but their mother was born abroad, their ‘country of origin’ was defined as the mother’s country of birth. If only the father was born abroad, their father’s country of birth was the person’s ‘country of origin’. Countries of origin were categorized into: The Netherlands, Europe excluding the Netherlands, Turkey, Morocco, Surinam, Dutch Caribbean, Indonesia, Other Africa, Other Asia, Other America and Oceania, and Unknown.

### Medical risk

Dutch national healthcare registry data were used to define underlying medical conditions that are associated with medical risk requiring vaccination prioritization. The national registry data comprised data from an all-payer claims database of (hospital-based) outpatient specialist care utilization, and data of medication at ATC-4 code level in the Netherlands (insurance covered medication dispensed outside hospitals and nursing homes). The all-payer claims database is managed by Vektis Health Care Information Center, and the medication dispensing database is managed by the National Health Care Institute (12, 13).

Supplementary file 2 lists the specific conditions that fall under two types of medical risk groups (i.e., high and intermediate medical risk), and shows the decisions that were made within the registry datasets to approximate these conditions. The medical high-risk group was defined by the Health Council of the Netherlands, based on conditions associated with high risk for severe COVID-19 (14). These patients were prioritized for primary series vaccination in 2021. The medical intermediate-risk group was based on the eligibility criteria for influenza vaccination in the Netherlands, and was also prioritized in the COVID-19 vaccination programme after the high-risk group (14). For both groups, age and long-term care use are part of the criteria, but these variables were excluded from the definition of medical risk as they were already included as time-varying variables. Medical risk was included as a categorical covariate: high-, intermediate-, or low-risk, with low-risk defined as not meeting the criteria for high-risk nor for intermediate-risk.

### Long-term care use

Start and end dates and types of LTC use (partly) reimbursed by insurance were available at individual level. Person-time was stratified according to LTC due to physical, sensory or mental disabilities (disability care, DC), and LTC for people aged 70 years or older. Long-term elderly care was further stratified into two broad groups, based on the level of care needs (i.e. ‘care profiles’ within the Dutch LTC system): LTC low 70+ (profiles 1-4) and LTC high 70+ (profiles 5-10). While LTC for elderly people with certain care profiles can be given either at home or in institutions (if feasible), the LTC low 70+ group was most likely to have received care at home and the LTC high 70+ group was most likely to have received institutional care, i.e., in a nursing home. This variable was time-dependent as LTC use could commence or end during the study period.

### Statistical analysis

VE was estimated using Cox proportional hazards regression with calendar time as underlying time scale and vaccination status as time-dependent exposure and unvaccinated person-time as reference. The outcome was death from COVID-19. Every person’s follow-up time started on 1 January 2021 and ended on date of death, date of emigration or 31 January 2022, whichever came first. Persons emigrating or with non-COVID-19 death were censored. VE was calculated as (1-hazard ratio (HR)) *100%. Vaccination statuses “Primary series – completed” and “Boosted” were stratified into months since vaccination. The proportional hazards assumption was met, as per visualization of Schoenfeld residuals over calendar time using R package ‘ggsurvminer’ v0.4.9 (15). VE analyses were repeated with all-cause mortality as secondary outcome.

The hazard ratio for death in the weeks following vaccination was estimated using similar models as the VE analysis, the only differences being the definition of the exposure status (risk periods after vaccination), the outcome (non-COVID-19 deaths), and censoring for COVID-19 death.

All analyses were stratified based on LTC use and birth year into the following strata: LTC high 70+ (born before 1951), LTC low 70+ (born before 1951), disability care (DC, all ages), other persons born before 1931 (90+ years), other persons born between 1931 and 1950 (70-89 years), other persons born between 1951 and 1970 (50-69 years), and other persons born between 1971 and 2009 (12-49 years). Sex, year of birth (as natural cubic spline with 4 degrees of freedom), medical risk (low, intermediate or high) and country of origin (in 10 categories) were included as covariates in the models. In tables in Supplementary File 1, estimates are also shown as crude and as partly adjusted estimates (adjusted for sex, year of birth and country of origin).

As per the CBS guidelines to prevent disclosure of groups or persons, all numbers smaller than 10 and incidences based on less than 10 observations are not presented, VE estimates higher than 99% are replaced by “>99%” and HR estimates lower than 0.01 are replaced by “<0.01”.

## Results

### VE against COVID-19 mortality

Table 1 shows characteristics of the study population, stratified into persons who survived the study period, persons who died of COVID-19, persons who died of other causes, and persons with a date of death but with a missing cause of death. Because vaccination status and LTC use are time-dependent, they are not included in Table 1. Prevalence of intermediate and high medical risk was much higher in the population that died during the study period (67.1% and 57.7% intermediate-risk, 8.9% and 15.1% high-risk) compared to those who survived (20.4% intermediate, 2.2% high risk). Older people, men, persons with a country of origin outside of the Netherlands, and those with an intermediate medical risk were most overrepresented in the population that died of COVID-19. Prevalence of high medical risk was highest among the population that died of causes other than COVID-19.

**Table 1.**
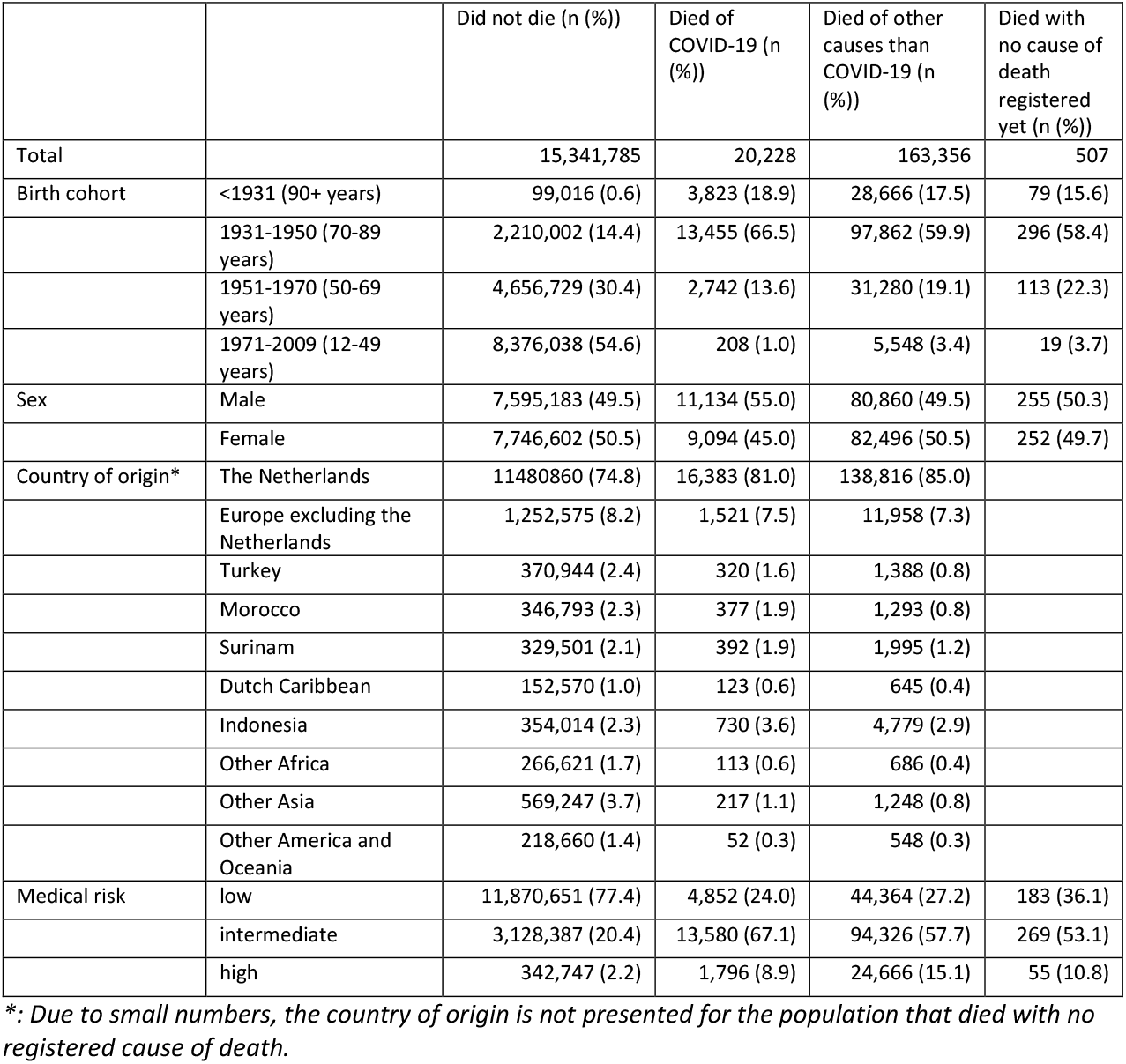
Study population characteristics, per outcome

|  |  | Did not die (n (%)) | Died of COVID-19 (n (%)) | Died of other causes than COVID-19 (n (%)) | Died with no cause of death registered yet (n (%)) |
| --- | --- | --- | --- | --- | --- |
| Total |  | 15,341,785 | 20,228 | 163,356 | 507 |
| Birth cohort | <1931 (90+ years) | 99,016 (0.6) | 3,823 (18.9) | 28,666 (17.5) | 79 (15.6) |
|  | 1931-1950 (70-89 years) | 2,210,002 (14.4) | 13,455 (66.5) | 97,862 (59.9) | 296 (58.4) |
|  | 1951-1970 (50-69 years) | 4,656,729 (30.4) | 2,742 (13.6) | 31,280 (19.1) | 113 (22.3) |
|  | 1971-2009 (12-49 years) | 8,376,038 (54.6) | 208 (1.0) | 5,548 (3.4) | 19 (3.7) |
| Sex | Male | 7,595,183 (49.5) | 11,134 (55.0) | 80,860 (49.5) | 255 (50.3) |
|  | Female | 7,746,602 (50.5) | 9,094 (45.0) | 82,496 (50.5) | 252 (49.7) |
| Country of origin* | The Netherlands | 11480860 (74.8) | 16,383 (81.0) | 138,816 (85.0) |  |
|  | Europe excluding the Netherlands | 1,252,575 (8.2) | 1,521 (7.5) | 11,958 (7.3) |  |
|  | Turkey | 370,944 (2.4) | 320 (1.6) | 1,388 (0.8) |  |
|  | Morocco | 346,793 (2.3) | 377 (1.9) | 1,293 (0.8) |  |
|  | Surinam | 329,501 (2.1) | 392 (1.9) | 1,995 (1.2) |  |
|  | Dutch Caribbean | 152,570 (1.0) | 123 (0.6) | 645 (0.4) |  |
|  | Indonesia | 354,014 (2.3) | 730 (3.6) | 4,779 (2.9) |  |
|  | Other Africa | 266,621 (1.7) | 113 (0.6) | 686 (0.4) |  |
|  | Other Asia | 569,247 (3.7) | 217 (1.1) | 1,248 (0.8) |  |
|  | Other America and Oceania | 218,660 (1.4) | 52 (0.3) | 548 (0.3) |  |
| Medical risk | low | 11,870,651 (77.4) | 4,852 (24.0) | 44,364 (27.2) | 183 (36.1) |
|  | intermediate | 3,128,387 (20.4) | 13,580 (67.1) | 94,326 (57.7) | 269 (53.1) |
|  | high | 342,747 (2.2) | 1,796 (8.9) | 24,666 (15.1) | 55 (10.8) |

\*: Due to small numbers, the country of origin is not presented for the population that died with no registered cause of death.

Figure 1 shows, per stratum of LTC use and birth cohort, the 4-week moving average of COVID-19 deaths per week (A) and crude incidence per 100,000 person-days (B), by vaccination status. The number and incidence of COVID-19 deaths was lowest in the 12-49 years age group and in people receiving disability care (DC). Throughout the study period, the incidence of COVID-19 deaths was highest in unvaccinated elderly. Figure 2 shows, per stratum of LTC use and birth cohort, the estimates of VE against COVID-19 mortality, which were very high (>90%) in the first two months after completion of the primary series. VE decreased gradually to around 75-80% at 6 months post-primary series. However, for the group receiving a high level of LTC aged 70 years or older, VE decreased to 66.5% at 6 months after completion of the primary series. VE estimates for 8, 9 and 10 months post-primary series decreased more rapidly, but these were based on a small selection of person-days as most people had received a booster vaccination before that time. For the group born between 1971 and 2009 (12-49 years old), the models did not converge for 6 or more months after primary vaccination due to low numbers of COVID-19 deaths. In this group, VE remained >90% up to 5 months post-primary vaccination. In the first two months after booster vaccination, VE was high in most groups (>93%); in the 90+ and 70+ high LTC groups the booster VE was >85%. Figure S2 shows the results of the secondary VE analysis, with all-cause mortality as outcome. This figure shows the same trends as Figure 1, but with lower VE, as COVID-19 comprised a minority of all deaths in the study period.

**Figure 1.**
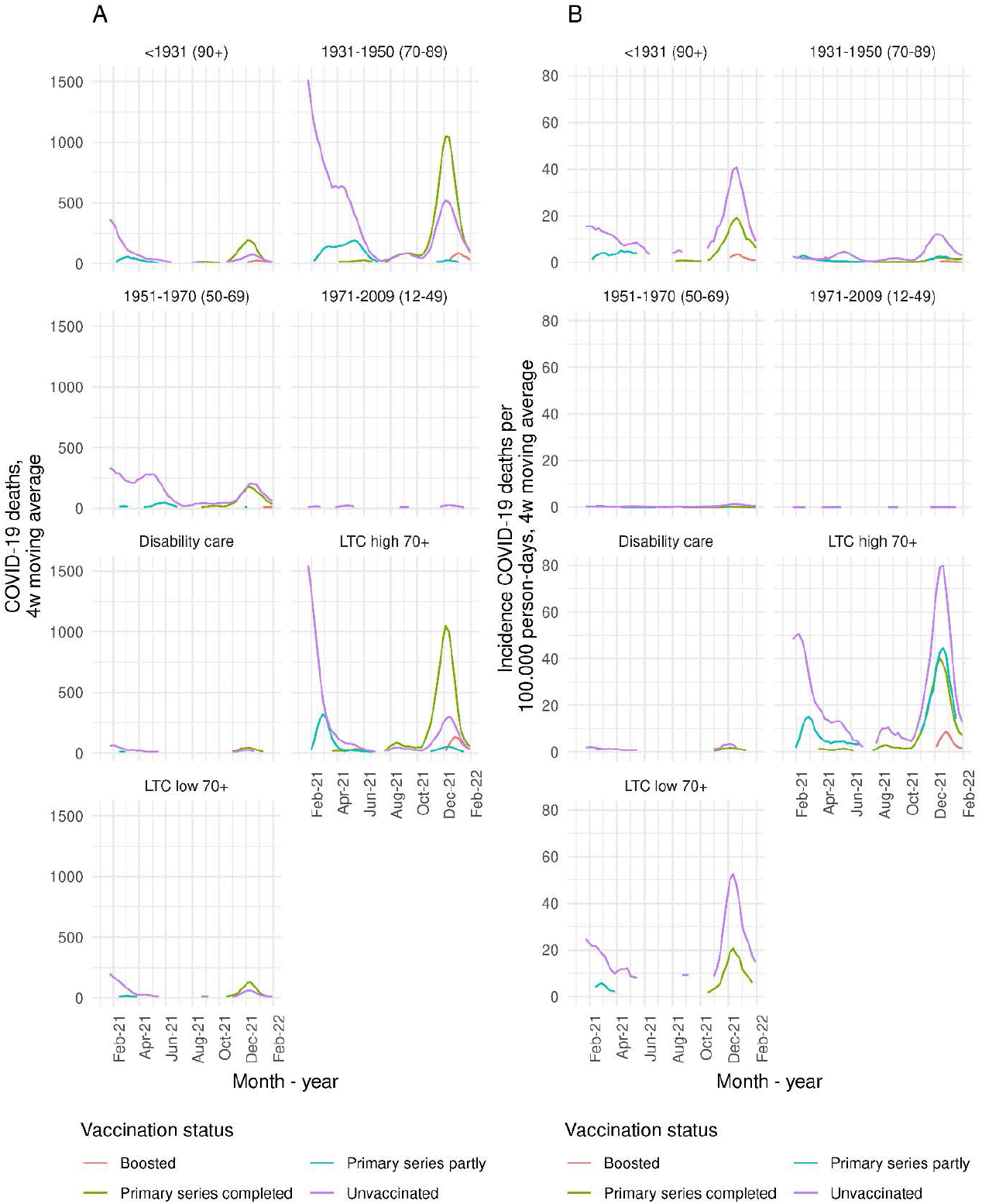
Four-week moving average of number of COVID-19 deaths per week (A) and incidence per 100.000 person-days (B), per stratum of long-term care use and birth cohort. Data points based on fewer than 10 deaths are not shown to prevent disclosure of groups or persons. A version of this Figure with varying y axes is Supplementary Figure S1.

**Figure 2.**
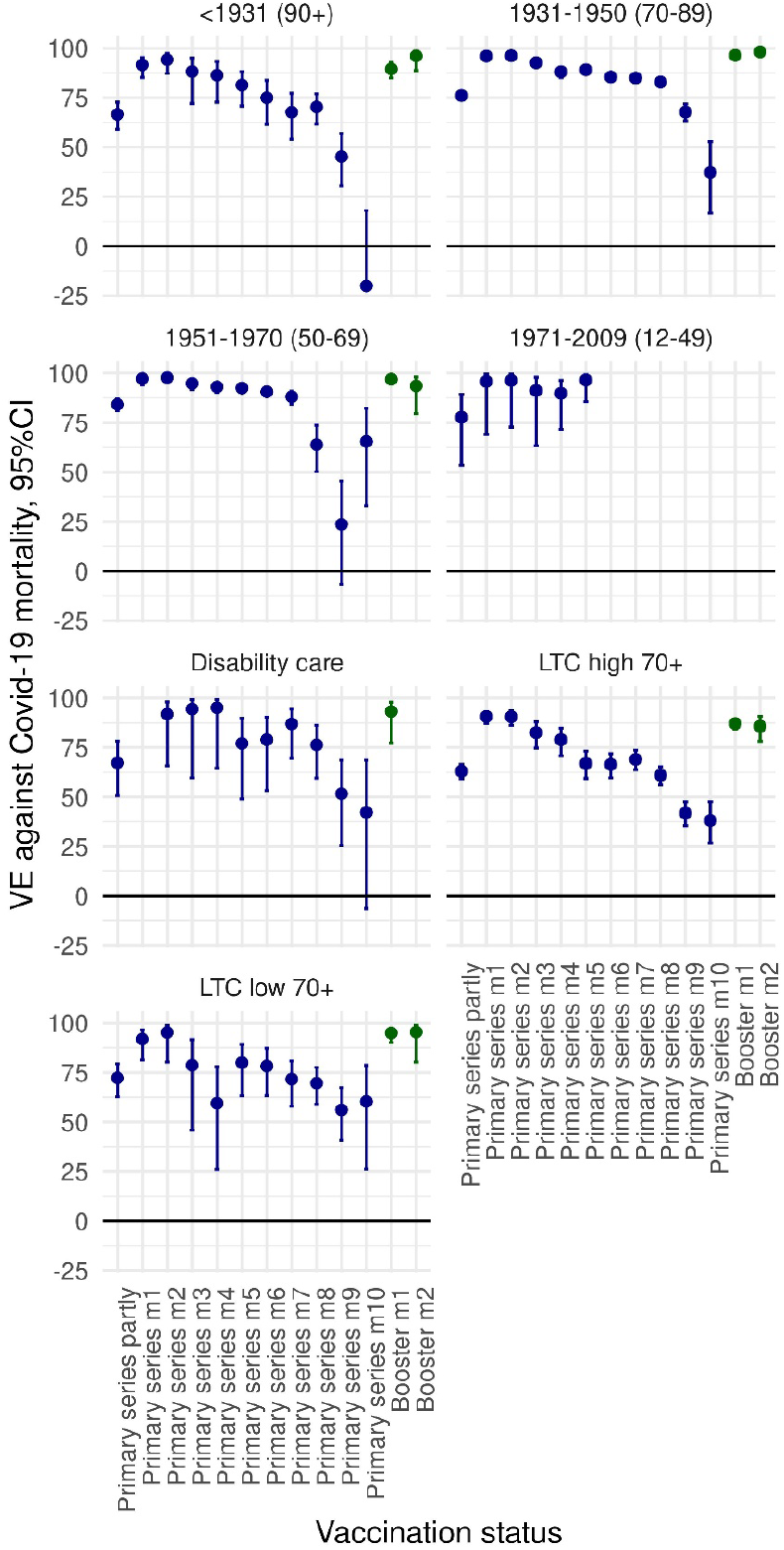
COVID-19 primary series and booster vaccine effectiveness (VE) estimates against COVID-19 mortality, compared to unvaccinated, with 95% confidence interval, per stratum of long-term care (LTC) use and birth cohort, adjusted for sex, year of birth, medical risk and country of origin. VE >99% are shown in grey without confidence interval. Underlying numbers, as well as crude and partly adjusted estimates, are shown in Supplementary Table S1.

### Risk of non-COVID-19 mortality after vaccination

Figure 3 shows, per stratum of LTC use and birth cohort, the 4-week moving average of non-COVID-19 deaths per week (A) and crude incidence per 100.000 person-days (B), per vaccination status. From the moment a group became eligible for vaccination, the absolute number of non-COVID deaths was soon highest among vaccinated persons because of the high coverage (earlier in 2021 for the oldest ages and LTC recipients, and mid-2021 for the youngest age groups). The incidence of non-COVID-19 deaths per 100.000 person-days was highest in unvaccinated individuals for most of the study period. For most groups, peaks in the incidence of non-COVID-19 mortality among vaccinated persons are visible at the start of the vaccination campaign (Figure S3). As panel A shows, these are based on low numbers, and these may possibly be explained by the prioritization of the most vulnerable persons. Also at the end of the study period, when the booster campaign started, the incidence of non-COVID-19 mortality in those having received the primary series increases in these age groups. Possibly, this reflects forgoing a booster dose by persons with a low remaining life expectancy.

**Figure 3.**
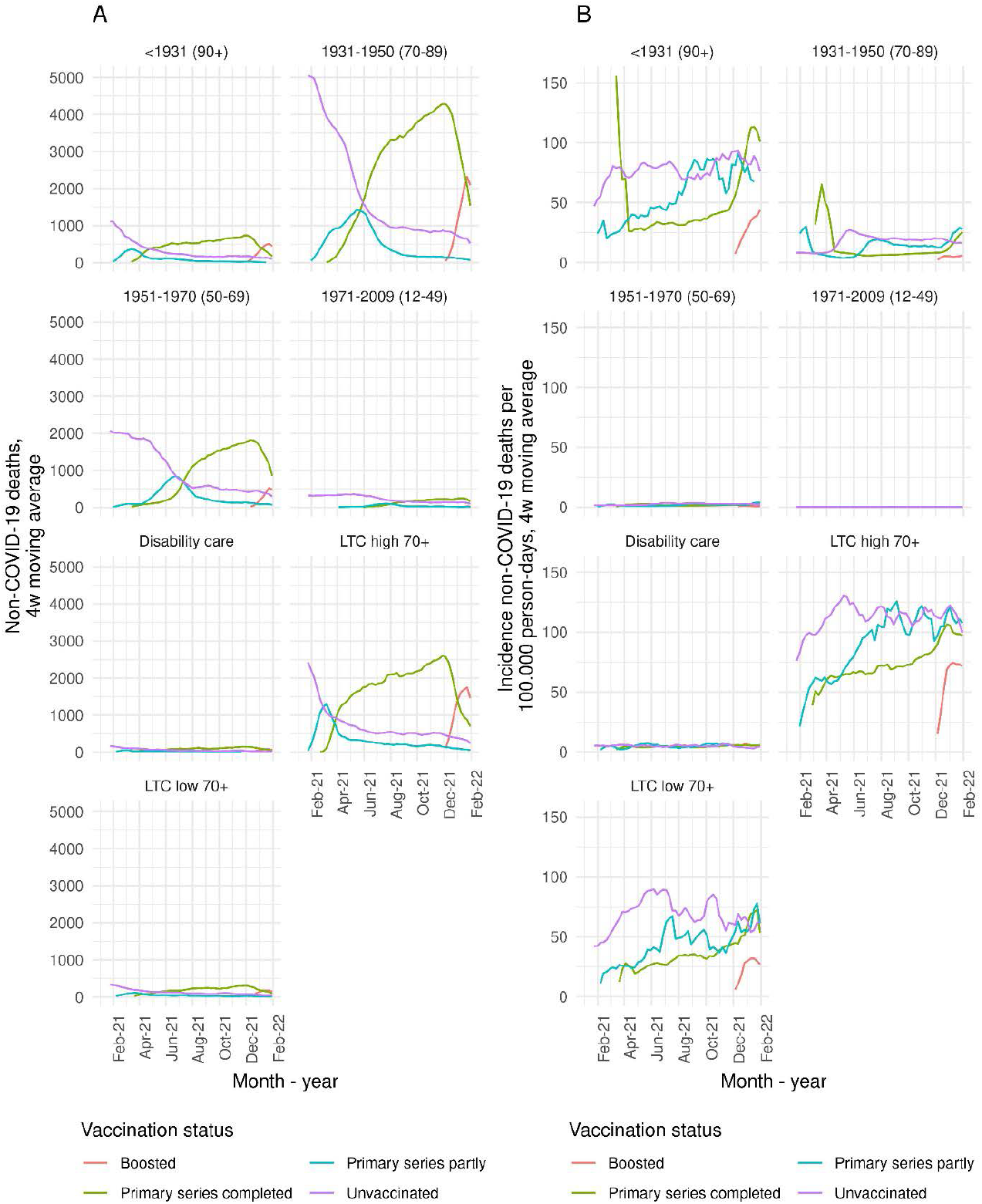
Four-week moving average of number of non-COVID-19 deaths per week (A) and incidence per 100.000 person-days (B), per stratum of long-term care use and birth cohort. Data points based on fewer than 10 deaths are not shown to prevent disclosure of groups or persons. A version of this Figure with varying y axes is Supplementary Figure S3.

Figure 4 shows, per stratum of LTC use and birth cohort, the HR estimates for risk of non-COVID death in the 5 weeks following a first dose of mRNA vaccine (A) and the 8 weeks following a first dose of vector vaccine (B), compared to unvaccinated person-time. For the first mRNA vaccination, nearly all estimates were significantly lower than 1, consistent with a reduced risk of non-COVID-19 death in these 5 weeks. After the first dose of vector vaccine, most HR estimates were also below 1, although these estimates had broader confidence intervals as these vaccines were not used as much in the Netherlands, except for the group of 60-64-year-olds who were specifically offered Vaxzevria initially. Figure 5 shows the HR estimates for risk of non-COVID-19 death following a second dose in the primary series, using person-time at least 2 weeks after a first dose as reference. Similarly, most estimates were below 1, except for the youngest age groups where after 4 weeks post-second dose the HR approached 1, consistent with no association between vaccination and non-COVID-deaths. Likewise, in the 8 weeks following booster vaccination, compared to person-time at least 3 months post-second dose, all HR estimates were below 1 (Figure 6).

**Figure 4.**
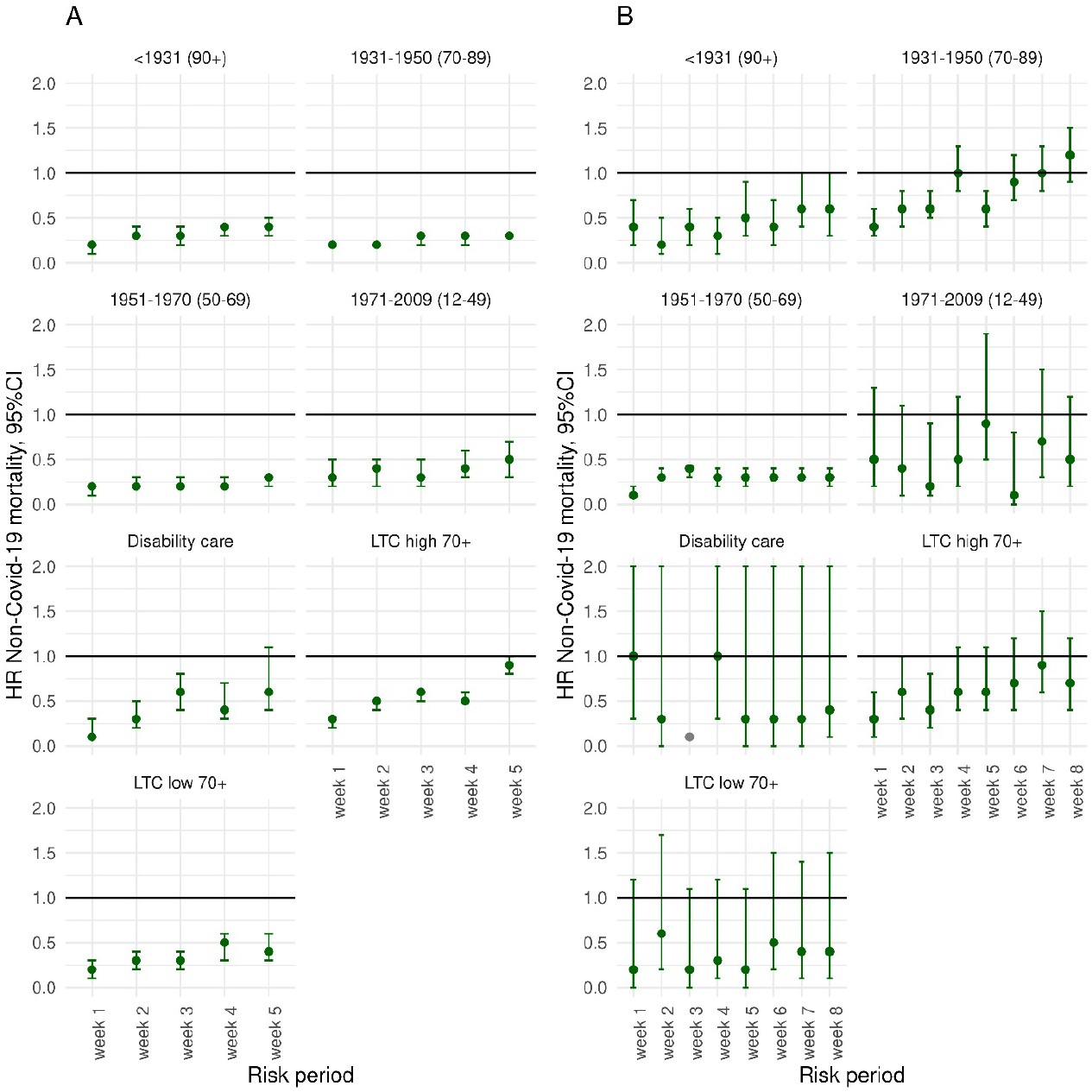
Hazard ratio (HR), with 95% confidence interval, for risk of non-COVID-19 mortality after a first dose of mRNA (A) or vector (B) vaccine compared to unvaccinated, per stratum of long-term care use and birth cohort, adjusted for sex, year of birth, medical risk and country of origin. HR <0.01 are shown in grey without confidence interval. Underlying numbers, as well as crude and partly adjusted estimates, are shown in Supplementary Table S3-4.

**Figure 5.**
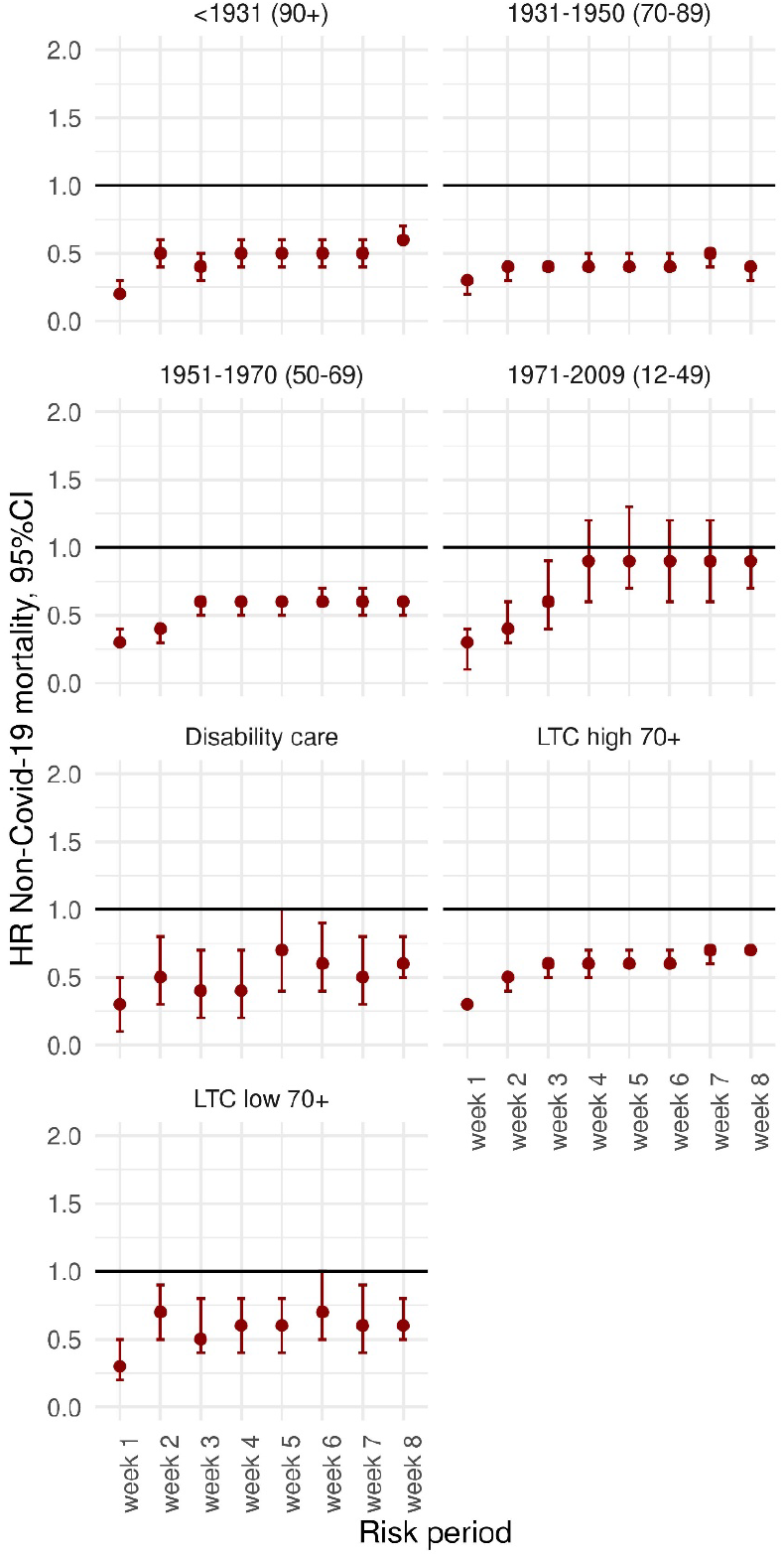
Hazard ratio (HR), with 95% confidence interval, for risk of non-COVID-19 mortality after a second dose of COVID-19 vaccine, compared to person-time at least 2 weeks post-first dose, per stratum of long-term care use and birth cohort, adjusted for sex, year of birth, medical risk and country of origin. Underlying numbers, as well as crude and partly adjusted estimates, are shown in Supplementary Table S5.

**Figure 6.**
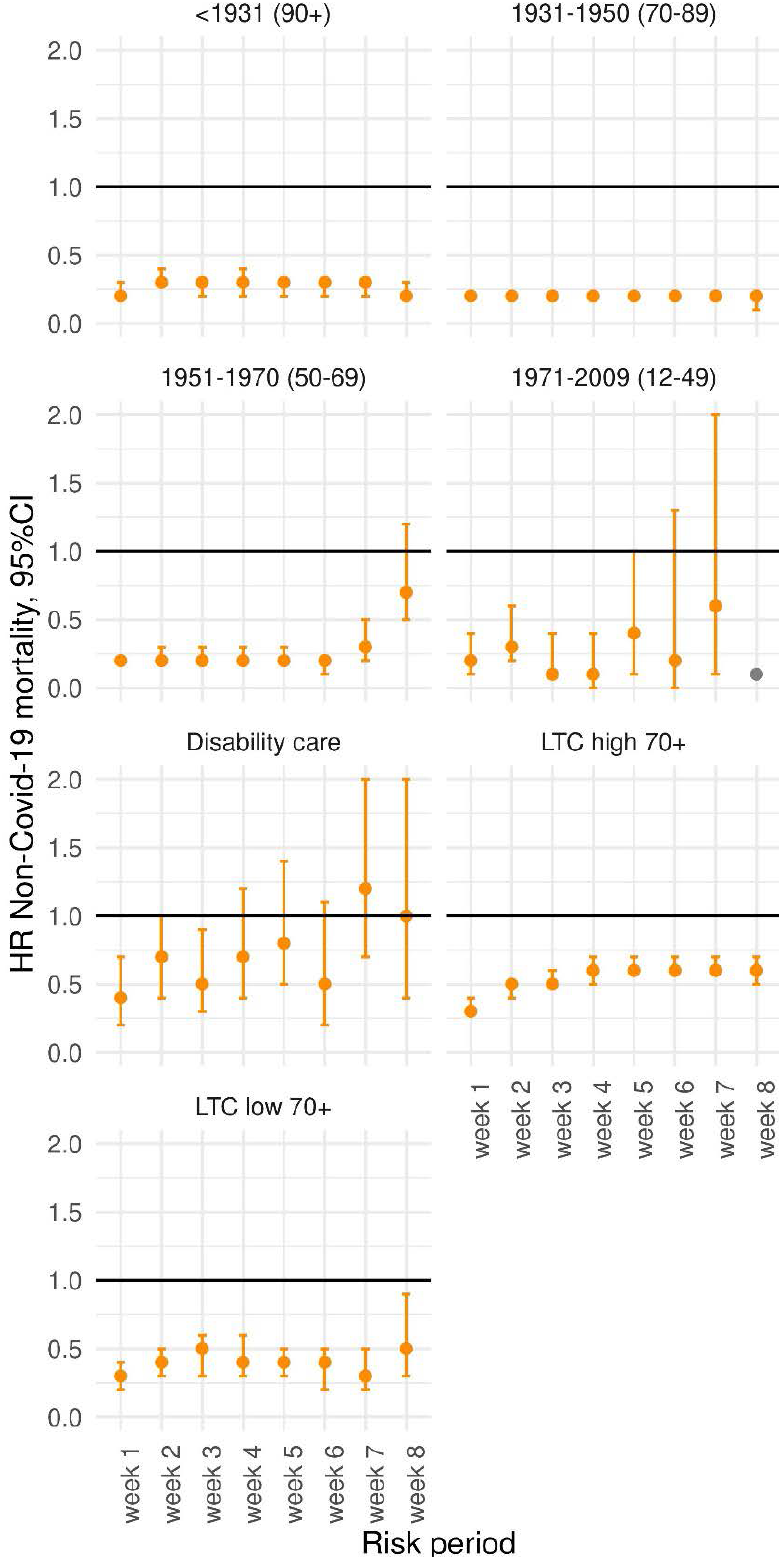
Hazard ratio (HR), with 95% confidence interval, for risk of non-COVID-19 mortality after a booster dose of COVID-19 vaccine, compared to person-time at least 3 months post-second dose, per stratum of long-term care use and birth cohort, adjusted for sex, year of birth, medical risk and country of origin. HR <0.01 are shown in grey without confidence interval. Underlying numbers, as well as crude and partly adjusted estimates, are shown in Supplementary Table S6.

## Discussion

Our results suggest that during January 2021-January 2022, VE against COVID-19 mortality was high in the Netherlands. While VE started at a very high level in all age groups in the two months after completion of the primary series, effectiveness decreased gradually in the months thereafter.

Booster vaccination restored VE, but with data up to 31 January 2022 we were not able to estimate VE three or more months after booster vaccination. The lower initial VE and faster waning among the oldest group and among elderly receiving a high level of LTC supports the prioritization of these groups within COVID-19 (re-)vaccination programmes. Our results are in line with other studies of VE against COVID-19 mortality (1, 2, 3, 4, 5, 6), that also found a very high VE shortly after primary or booster vaccination, and faster VE waning among LTC recipients (3).

Our analyses of risk of non-COVID-19 mortality in the 5 weeks (after mRNA dose 1) or 8 weeks (after other doses) following vaccination showed a lower to comparable risk, compared to the vaccination status before the respective vaccine dose. This was observed for all ages and including LTC recipients. Consistent with our results, previous studies also found lower non-COVID-19 mortality rates after COVID-19 vaccination (6, 16, 17). A true protective effect of COVID-19 vaccination on non-COVID-19 mortality is biologically implausible. Likely, healthy vaccinee bias has affected the results (18). For example, fever was a contra-indication for vaccination, resulting in a selection of relatively healthy person-time shortly after vaccination. Also, people with a short remaining life expectancy may have opted to forego vaccination. Another possibility is that in deaths that were not attributed to COVID-19, SARS-CoV-2 infection actually had an unrecognized role in the causal pathway to death.

We did not stratify the study period corresponding to dominant SARS-CoV-2 variant. At the start of 2021, the SARS-CoV-2 wildtype still dominated, which was replaced by the Alpha variant in Spring 2021. During June 2021, Delta replaced Alpha, and around the turn of the year, Omicron BA.1 replaced Delta. Stratification into variant periods was explored, but did not result in informative estimates due to small numbers. Also, because the booster campaign coincided with the emergence of Omicron BA.1, the booster VE could not be stratified for different variants. We do not expect large differences in VE against COVID-19 mortality between Alpha and Delta, because the VE against hospitalization was similar for these variants (19, 20) A study from Italy found similar VE against COVID-19 mortality for the Alpha and Delta era’s in the first 3 months post-vaccination (21). However, it is possible that the lower VE estimates in months 8 to 10 post-primary series are associated with the emergence of Omicron BA.1 in the final months of the study period, consistent with a sharp decrease in the VE against hospitalization with Omicron compared to Delta for the primary series (22, 23).

The data we used has limitations. Because national registration of vaccination in CIMS is dependent on informed consent of the vaccinee, this register is incomplete by design. For the VE analysis this means that a part of the population defined as unvaccinated in the analysis was in fact vaccinated. Of people vaccinated at the Municipal Health Services (around 86% of all primary series in the Netherlands (24)), around 7% of primary series recipients did not give consent for registration in CIMS, and around 5% of booster vaccinations were was not registered in CIMS. The resulting misclassification will have led to an underestimation of VE. A separate study by our group aimed to quantify the possible bias in VE due to this misclassification, and found that a non-consent rate of 7%, combined with a vaccination coverage of 90% and a VE of 90% will result in an underestimation of VE of around 5 percentage points (25). As this bias is smaller with lower vaccination coverage, underestimation of VE is likely smaller for younger age groups. Similarly, for the analysis of non-COVID-19 mortality the vaccination effect might be diluted due to this misclassification. Likely the bias due to misclassification is smaller for the analyses of the second dose compared to one dose and the booster compared to two doses, if vaccinees giving consent for central registration for the first vaccine dose will also do so for subsequent doses.

Another important limitation is lack of data on previous SARS-CoV-2 infections in the study population. According to a Dutch seroprevalence study, in February/March 2021 around 12% of the population had experienced a SARS-CoV-2 infection, increasing to around 25% in November/December 2021 and to 60% in March/April 2022 (26). This will have contributed to protection against death from subsequent infections. Unvaccinated populations likely accrued infections sooner, more often and more severe, leading to a faster build-up of infection-induced immunity, which may lead to lower VE estimates over time without actual waning of VE being present (27). We plan to account for this in future analyses, including data on confirmed infections, which was not available at the time of this study. The lack of data on infections is less likely to influence the analyses of non-COVID-19 mortality. However, infection data could be useful to assess the effects of infections, by for example re-classifying any death within 30 days after a positive SARS-CoV-2 test as COVID-19 mortality in a sensitivity analysis.

The data we used to define medical risk groups also have important limitations. Both the medication dispensing data, and the registration data of outpatient specialist care use, were most recently available for 2020. Therefore, persons with high or intermediate risk who first visited the medical specialist in 2021 will have been misclassified as low-risk and also persons that did not visit the medical specialist yearly could have been misclassified as low-risk. For instance, the specialist care use data was not recent enough to identify all persons with hematological malignancies diagnosed in the last five years, a subgroup of the medical high-risk group as defined by the Health Council of the Netherlands, as we could only identify persons that visited the medical specialist between 2016 and 2020. Although most risk groups could be indicated well with the registration data of specialist care use, not all relevant conditions could be identified. For instance, an unknown proportion of the risk group of people with morbid obesity might receive care from the general physician only, and if referred to the medical specialist the specific diagnostic code for morbid obesity might not always be used (e.g., if the specialist care is needed for an associated comorbid condition). In our analyses, adjustment for medical risk increased VE estimates (see Supplementary Tables S1 and S2). Before adjustment, VE against all-cause mortality was negative for some groups at 7 or more months post-primary series. After adjustment, VE estimates increased substantially, consistent with our hypothesis that negative VE was caused by an overrepresentation of individuals with comorbidity in the later months post-vaccination. Contrary, estimates of the HR of non-COVID-19 mortality shortly after vaccination showed very little changes after adjustment for medical risk group, indicating that chronic conditions hardly contributed to a presumed healthy vaccinee bias in these estimates (see Supplementary Tables S3-S6). Likely, more acute conditions such as fever played a larger role in a healthy vaccinee bias for these estimates.

Aside from the data limitations, the observational nature of this study warrants caution when interpreting its results. Characteristics associated with uptake of vaccination, and with consent for central registration, may also be associated with the chance of SARS-CoV-2 exposure or other health-related behaviors. The direction of such effects is unclear: on the one hand, vaccinated people may exhibit more health-conscious behavior and lower their risk of SARS-CoV-2 exposure, on the other hand risk perception may be lowered after vaccination, leading to more exposure. Policies such as vaccination or test requirements for entry into establishments or events could have increased the latter effect.

In conclusion, we found high effectiveness of COVID-19 vaccination against COVID-19 mortality, and no indication of increased risk of non-COVID mortality after vaccination. Our study confirms the major public health benefits of primary and booster COVID-19 vaccinations, especially among vulnerable populations (28).

## Data Availability

Under certain conditions, these microdata are accessible for statistical and scientific
research. For further information: microdata -at- cbs.nl.

## References

1. Haas EJ, Angulo FJ, McLaughlin JM, Anis E, Singer SR, Khan F, et al. Impact and effectiveness of mRNA BNT162b2 vaccine against SARS-CoV-2 infections and COVID-19 cases, hospitalisations, and deaths following a nationwide vaccination campaign in Israel: an observational study using national surveillance data. Lancet. 2021;397(10287):1819–29.

2. Andrews N, Tessier E, Stowe J, Gower C, Kirsebom F, Simmons R, et al. Duration of Protection against Mild and Severe Disease by Covid-19 Vaccines. N Engl J Med. 2022;386(4):340–50.

3. Shrotri M, Krutikov M, Palmer T, Giddings R, Azmi B, Subbarao S, et al. Vaccine effectiveness of the first dose of ChAdOx1 nCoV-19 and BNT162b2 against SARS-CoV-2 infection in residents of long-term care facilities in England (VIVALDI): a prospective cohort study. Lancet Infect Dis. 2021;21(11):1529–38.

4. Nunes B, Rodrigues AP, Kislaya I, Cruz C, Peralta-Santos A, Lima J, et al. mRNA vaccine effectiveness against COVID-19-related hospitalisations and deaths in older adults: a cohort study based on data linkage of national health registries in Portugal, February to August 2021. Euro Surveill. 2021;26(38).

5. Lin DY, Gu Y, Xu Y, Wheeler B, Young H, Sunny SK, et al. Association of Primary and Booster Vaccination and Prior Infection With SARS-CoV-2 Infection and Severe COVID-19 Outcomes. JAMA. 2022;328(14):1415–26.

6. Horne EMF, Hulme WJ, Keogh RH, Palmer TM, Williamson EJ, Parker EPK, et al. Waning effectiveness of BNT162b2 and ChAdOx1 covid-19 vaccines over six months since second dose: OpenSAFELY cohort study using linked electronic health records. BMJ. 2022;378:e071249.

7. European Medicines Agency, 2022. https://www.ema.europa.eu/en/human-regulatory/overview/public-health-threats/coronavirus-disease-covid-19/treatments-vaccines/vaccines-covid-19/safety-covid-19-vaccines#latest-safety-information-section.

8. Hafeez MU, Ikram M, Shafiq Z, Sarfraz A, Sarfraz Z, Jaiswal V, et al. COVID-19 Vaccine-Associated Thrombosis With Thrombocytopenia Syndrome (TTS): A Systematic Review and Post Hoc Analysis. Clin Appl Thromb Hemost. 2021;27:10760296211048815.

9. Andrews NJ, Stowe J, Ramsay ME, Miller E. Risk of venous thrombotic events and thrombocytopenia in sequential time periods after ChAdOx1 and BNT162b2 COVID-19 vaccines: A national cohort study in England. Lancet Reg Health Eur. 2022;13:100260.

10. Proof of vaccination: requirements and validity: Government of the Netherlands; [cited 2023 22 May]. Available from: https://www.government.nl/topics/coronavirus-covid-19/covid-certificate/proof-of-vaccination/requirements-and-validity.

11. Patalon T, Gazit S, Pitzer VE, Prunas O, Warren JL, Weinberger DM. Odds of Testing Positive for SARS-CoV-2 Following Receipt of 3 vs 2 Doses of the BNT162b2 mRNA Vaccine. JAMA Intern Med. 2022;182(2):179–84.

12. Netherlands) CS. Persons with dispensed medicines. 2022.

13. Geurten RJ, Struijs JN, Elissen AMJ, Bilo HJG, van Tilburg C, Ruwaard D. Delineating the Type 2 Diabetes Population in the Netherlands Using an All-Payer Claims Database: Specialist Care, Medication Utilization and Expenditures 2016-2018. Pharmacoecon Open. 2022;6(2):219–29.

14. RIVM. COVID-19-vaccinatie uitvoeringsrichtlijn. [COVID-19 vaccination implementing guideline] https://lci.rivm.nl/richtlijnen/covid-19-vaccinatie 2022.

15. Kassambara A, Kosinski M, Biecek P, Fabian S. survminer: Drawing Survival Curves using ‘ggplot2’ 2021 [Available from: https://CRAN.R[1]project.org/package=survminer.

16. Xu S, Huang R, Sy LS, Glenn SC, Ryan DS, Morrissette K, et al. COVID-19 Vaccination and Non-COVID-19 Mortality Risk - Seven Integrated Health Care Organizations, United States, December 14, 2020-July 31, 2021. MMWR Morb Mortal Wkly Rep. 2021;70(43):1520–4.

17. Xu S, Huang R, Sy LS, Hong V, Glenn SC, Ryan DS, et al. A safety study evaluating non-COVID-19 mortality risk following COVID-19 vaccination. Vaccine. 2023;41(3):844–54.

18. Remschmidt C, Wichmann O, Harder T. Frequency and impact of confounding by indication and healthy vaccinee bias in observational studies assessing influenza vaccine effectiveness: a systematic review. BMC Infect Dis. 2015;15:429.

19. Lauring AS, Tenforde MW, Chappell JD, Gaglani M, Ginde AA, McNeal T, et al. Clinical severity of, and effectiveness of mRNA vaccines against, covid-19 from omicron, delta, and alpha SARS-CoV-2 variants in the United States: prospective observational study. BMJ. 2022;376:e069761.

20. RIVM. Effectiviteit van COVID-19 vaccinatie tegen ziekenhuis-en intensive-care opname in Nederland. [Effectiveness of COVID-19 vaccination against hospitalization and ICU admission in the Netherlands, in Dutch] https://www.rivm.nl/documenten/studie-effectiviteit-van-covid-19-vaccinatie-tegen-ziekenhuis-en-intensive-care-opname. 2021.

21. Homan T, Mazzilli S, Chieti A, Musa A, Roth A, Fortunato F, et al. Covid-19 vaccination programme effectiveness against SARS-CoV-2 related infections, hospital admissions and deaths in the Apulia region of Italy: a one-year retrospective cohort study. Sci Rep. 2022;12(1):18597.

22. RIVM. Effectiviteit van COVID-19 vaccinatie tegen ziekenhuis-en intensive-care opname in Nederland (opnames 1 december 2021 – 8 februari 2022). [Effectiveness of COVID-19 vaccination against hospitalization and ICU admission in the Netherlands, 1 December 2021 – 8 February 2022, in Dutch] https://www.rivm.nl/documenten/effectiviteit-van-covid-19-vaccinatie-tegen-ziekenhuis-en-intensive-care-opname-in-6. 2022.

23. Tenforde MW, Self WH, Gaglani M, Ginde AA, Douin DJ, Talbot HK, et al. Effectiveness of mRNA Vaccination in Preventing COVID-19-Associated Invasive Mechanical Ventilation and Death - United States, March 2021-January 2022. MMWR Morb Mortal Wkly Rep. 2022;71(12):459–65.

24. RIVM. Vaccination figures: 6 January – 26 December 2021. https://www.rivm.nl/en/covid-19-vaccination/archive-covid-19-vaccination-figures-2021. 2021.

25. Van Werkhoven CH, De Gier B, McDonald SA, De Melker HE, Hahne SJM, Van den Hof S, et al. Information bias of vaccine effectiveness estimation due to informed consent for national registration of COVID-19 vaccination: estimation and correction using a data augmentation model. medRxiv. 2023:2023.05.23.23290384.

26. RIVM. Resultaten Pienter Corona studie ronde 7 [in Dutch] https://www.rivm.nl/pienter-corona-onderzoek/resultaten. 2022.

27. Kahn R, Schrag SJ, Verani JR, Lipsitch M. Identifying and Alleviating Bias Due to Differential Depletion of Susceptible People in Postmarketing Evaluations of COVID-19 Vaccines. Am J Epidemiol. 2022;191(5):800–11.

28. Mesle MM, Brown J, Mook P, Hagan J, Pastore R, Bundle N, et al. Estimated number of deaths directly averted in people 60 years and older as a result of COVID-19 vaccination in the WHO European Region, December 2020 to November 2021. Euro Surveill. 2021;26(47).

