## Supplementary material for "Effect of COVID-19 vaccination on mortality by COVID-19 and on mortality by other causes, the Netherlands, January 2021- January 2022": Table S

### Supplementary file 1 to Effect of COVID-19 vaccination on mortality by COVID-19 and on mortality by other causes, the Netherlands, January 2021- January 2022

Table S1. VE estimates of primary and booster vaccination against COVID-19 mortality, with 95% confidence interval, per stratum of long-term care use and birth cohort, crude, adjusted for sex, year of birth and country of origin (adjusted 1) and adjusted for sex, year of birth, country of origin and medical risk (adjusted 2). Numbers below 10 are not shown to prevent disclosure of groups or persons.

| Stratum | Vaccination status | Person-time (x 1000 days) | COVID-19 deaths | VE (95% CI) crude | VE (95% CI) adjusted 1 | VE (95% CI) adjusted 2 |
|---|---|---|---|---|---|---|
| **LTC high 70+** | Unvaccinated | 10778 | 3282 | [ref] | [ref] | [ref] |
| **LTC high 70+** | Primary series partly | 6368 | 589 | 62.3% (58.4-65.8) | 63.2% (59.4-66.6) | 63.1% (59.3-66.5) |
| **LTC high 70+** | Primary series m1 | 3361 | 42 | 90.4% (86.8-93) | 90.8% (87.3-93.3) | 90.7% (87.2-93.2) |
| **LTC high 70+** | Primary series m2 | 3291 | 31 | 90.4% (86-93.4) | 90.6% (86.3-93.6) | 90.6% (86.3-93.6) |
| **LTC high 70+** | Primary series m3 | 3338 | 34 | 82.8% (75.1-88.1) | 82.6% (74.8-88) | 82.6% (74.8-88) |
| **LTC high 70+** | Primary series m4 | 3311 | 41 | 79.1% (71-84.9) | 78.9% (70.8-84.8) | 79% (70.9-84.9) |
| **LTC high 70+** | Primary series m5 | 3368 | 125 | 67.6% (60.3-73.6) | 67% (59.5-73.1) | 67% (59.5-73.2) |
| **LTC high 70+** | Primary series m6 | 3310 | 151 | 68.5% (62.3-73.7) | 66.3% (59.7-71.9) | 66.5% (59.8-72) |
| **LTC high 70+** | Primary series m7 | 3113 | 198 | 69.2% (63.9-73.7) | 68.9% (63.5-73.5) | 69.1% (63.8-73.6) |
| **LTC high 70+** | Primary series m8 | 2779 | 468 | 58.5% (53.3-63.1) | 60.9% (56-65.3) | 61.1% (56.2-65.4) |
| **LTC high 70+** | Primary series m9 | 1917 | 793 | 38.7% (32-44.7) | 41.8% (35.4-47.6) | 41.9% (35.5-47.6) |
| **LTC high 70+** | Primary series m10 | 933 | 203 | 35.8% (24.2-45.6) | 38.2% (26.9-47.7) | 38.2% (26.9-47.7) |
| **LTC high 70+** | Booster m1 | 2629 | 151 | 86.5% (83.8-88.8) | 87.2% (84.6-89.4) | 87.2% (84.5-89.4) |
| **LTC high 70+** | Booster m2 | 1401 | 27 | 84.9% (76.8-90.2) | 85.8% (78.2-90.8) | 85.8% (78.2-90.8) |
| **LTC low 70+** | Unvaccinated | 3159 | 525 | [ref] | [ref] | [ref] |
| **LTC low 70+** | Primary series partly | 1556 | 54 | 71% (61.1-78.5) | 72.1% (62.5-79.3) | 72.3% (62.7-79.4) |
| **LTC low 70+** | Primary series m1 | 783 | <10 | 91.3% (80.1-96.2) | 91.9% (81.3-96.5) | 91.9% (81.4-96.5) |
| **LTC low 70+** | Primary series m2 | 767 | <10 | 95% (79.4-98.8) | 95.3% (80.3-98.9) | 95.3% (80.3-98.9) |
| **LTC low 70+** | Primary series m3 | 765 | <10 | 79.1% (47-91.8) | 78.6% (45.6-91.6) | 78.7% (45.9-91.7) |

| | | | | | | |
|---|---|---|---|---|---|---|
| **LTC low 70+** | Primary series m4 | 763 | 13 | 60% (26.8-78.1) | 58.8% (24.6-77.5) | 59.5% (25.9-77.9) |
| **LTC low 70+** | Primary series m5 | 764 | 12 | 79.9% (63.1-89.1) | 79.7% (62.6-89) | 80% (63.2-89.2) |
| **LTC low 70+** | Primary series m6 | 744 | 16 | 79.5% (65.5-87.9) | 77.9% (62.5-86.9) | 78.3% (63.3-87.2) |
| **LTC low 70+** | Primary series m7 | 702 | 32 | 72.5% (59.3-81.4) | 70.9% (56.8-80.4) | 71.7% (58-80.9) |
| **LTC low 70+** | Primary series m8 | 596 | 65 | 67.7% (56.3-76.1) | 69% (58.1-77.1) | 69.7% (59-77.6) |
| **LTC low 70+** | Primary series m9 | 324 | 71 | 51% (34.1-63.5) | 55.6% (40.3-67) | 56.2% (41-67.4) |
| **LTC low 70+** | Primary series m10 | 122 | 12 | 54.8% (16-75.7) | 60% (25.5-78.5) | 60.4% (26.2-78.7) |
| **LTC low 70+** | Booster m1 | 657 | 10 | 94.5% (89.2-97.1) | 94.9% (90.2-97.4) | 95% (90.2-97.4) |
| **LTC low 70+** | Booster m2 | 302 | <10 | 94.8% (77.5-98.8) | 95.4% (80.2-99) | 95.5% (80.3-99) |
| **Disability care** | Unvaccinated | 18128 | 206 | [ref] | [ref] | [ref] |
| **Disability care** | Primary series partly | 5744 | 28 | 47.8% (21.7-65.3) | 67.8% (51.4-78.6) | 67.3% (50.7-78.3) |
| **Disability care** | Primary series m1 | 2948 | <10 | >99% (--) | >99% (--) | >99% (--) |
| **Disability care** | Primary series m2 | 2895 | <10 | 86.3% (43.7-96.7) | 92.1% (66.8-98.1) | 91.9% (65.9-98.1) |
| **Disability care** | Primary series m3 | 2861 | <10 | 93.7% (54.6-99.1) | 94.6% (60.3-99.3) | 94.5% (59.8-99.3) |
| **Disability care** | Primary series m4 | 2827 | <10 | 94.9% (63.1-99.3) | 95% (63.6-99.3) | 95.2% (64.6-99.3) |
| **Disability care** | Primary series m5 | 2770 | <10 | 65.7% (24.4-84.4) | 76.5% (47.4-89.5) | 77.1% (49-89.8) |
| **Disability care** | Primary series m6 | 2468 | <10 | 58.1% (7.4-81.1) | 78.2% (51.5-90.2) | 79% (53.4-90.5) |
| **Disability care** | Primary series m7 | 2150 | <10 | 73.2% (37.2-88.5) | 86.7% (68.8-94.3) | 87% (69.7-94.5) |
| **Disability care** | Primary series m8 | 1913 | 18 | 49.5% (13.4-70.5) | 76.5% (59.7-86.3) | 76.4% (59.6-86.3) |
| **Disability care** | Primary series m9 | 1497 | 37 | -9.6% (-68.7-28.8) | 53.5% (28.5-69.8) | 51.7% (25.6-68.7) |
| **Disability care** | Primary series m10 | 794 | 15 | -46.7% (-168.9-20) | 45.7% (0.3-70.4) | 42.3% (-6.4-68.7) |
| **Disability care** | Booster m1 | 1918 | <10 | 82.1% (39.8-94.7) | 93.5% (78.2-98.1) | 93.3% (77.3-98) |
| **Disability care** | Booster m2 | 580 | <10 | >99% (--) | >99% (--) | >99% (--) |
| **<1931 (90+)** | Unvaccinated | 7375 | 899 | [ref] | [ref] | [ref] |
| **<1931 (90+)** | Primary series partly | 3857 | 129 | 67.1% (59.7-73.1) | 66.1% (58.5-72.3) | 66.6% (59.1-72.7) |
| **<1931 (90+)** | Primary series m1 | 2004 | 13 | 91.9% (85.6-95.4) | 91.5% (85-95.2) | 91.6% (85.2-95.3) |
| **<1931 (90+)** | Primary series m2 | 1936 | <10 | 94.3% (87.6-97.4) | 94.1% (87.1-97.3) | 94.2% (87.3-97.3) |
| **<1931 (90+)** | Primary series m3 | 1915 | <10 | 88.3% (72.3-95.1) | 87.9% (71.4-94.9) | 88.2% (72.1-95) |
| **<1931 (90+)** | Primary series m4 | 1853 | <10 | 86.3% (72.5-93.2) | 86% (72-93) | 86.4% (72.7-93.2) |

| | | | | | | |
|---|---|---|---|---|---|---|
| **<1931 (90+)** | Primary series m5 | 1835 | 24 | 81.2% (70.4-88) | 80.9% (69.9-87.8) | 81.4% (70.8-88.2) |
| **<1931 (90+)** | Primary series m6 | 1732 | 26 | 74.6% (60.8-83.6) | 74.1% (60-83.2) | 75% (61.5-83.8) |
| **<1931 (90+)** | Primary series m7 | 1567 | 45 | 67.3% (53.5-77.1) | 66.1% (51.7-76.2) | 67.7% (54-77.3) |
| **<1931 (90+)** | Primary series m8 | 1413 | 111 | 70.1% (61.4-76.8) | 68.9% (59.9-75.9) | 70.3% (61.6-77) |
| **<1931 (90+)** | Primary series m9 | 671 | 138 | 44.7% (29.8-56.4) | 42.8% (27.4-55) | 45.2% (30.4-56.9) |
| **<1931 (90+)** | Primary series m10 | 157 | 32 | -31.4% (-100.7-14) | -33.3% (-103.6-12.8) | -25.4% (-91.8-18) |
| **<1931 (90+)** | Booster m1 | 1462 | 40 | 89.4% (84.8-92.6) | 89.1% (84.3-92.4) | 89.5% (85-92.7) |
| **<1931 (90+)** | Booster m2 | 891 | <10 | 96% (88.6-98.6) | 95.9% (88.4-98.5) | 96.1% (88.8-98.6) |
| **1931-1950 (70-89)** | Unvaccinated | 250627 | 5591 | [ref] | [ref] | [ref] |
| **1931-1950 (70-89)** | Primary series partly | 108277 | 696 | 71.7% (68.9-74.2) | 75.3% (73-77.3) | 76.1% (73.9-78.1) |
| **1931-1950 (70-89)** | Primary series m1 | 59638 | 57 | 93.2% (91.1-94.8) | 95.9% (94.6-96.8) | 96% (94.8-97) |
| **1931-1950 (70-89)** | Primary series m2 | 59665 | 38 | 94.5% (92.3-96) | 96% (94.5-97.1) | 96.2% (94.8-97.3) |
| **1931-1950 (70-89)** | Primary series m3 | 59546 | 51 | 94.2% (92.2-95.7) | 92.2% (89.6-94.2) | 92.7% (90.3-94.6) |
| **1931-1950 (70-89)** | Primary series m4 | 58656 | 94 | 89.6% (87-91.7) | 87.2% (84-89.8) | 88.1% (85.2-90.5) |
| **1931-1950 (70-89)** | Primary series m5 | 58535 | 149 | 91.6% (90-93) | 88.1% (85.8-90.1) | 89.1% (86.9-90.8) |
| **1931-1950 (70-89)** | Primary series m6 | 57014 | 419 | 89.9% (88.7-91) | 84.2% (82.3-85.9) | 85.5% (83.7-87.1) |
| **1931-1950 (70-89)** | Primary series m7 | 46666 | 550 | 84.7% (83-86.2) | 83.6% (81.8-85.2) | 85% (83.4-86.4) |
| **1931-1950 (70-89)** | Primary series m8 | 20076 | 543 | 68.2% (64.7-71.3) | 81.1% (79.1-83) | 82.9% (81.1-84.6) |
| **1931-1950 (70-89)** | Primary series m9 | 4768 | 305 | 15.6% (3.8-25.9) | 63.2% (58-67.8) | 67.8% (63.3-71.8) |
| **1931-1950 (70-89)** | Primary series m10 | 801 | 56 | -99.4% (-163.9--50.6) | 18.9% (-7.7-38.9) | 37.3% (16.6-52.8) |
| **1931-1950 (70-89)** | Booster m1 | 48724 | 109 | 94.9% (93.8-95.9) | 96.3% (95.5-97) | 96.4% (95.6-97.1) |
| **1931-1950 (70-89)** | Booster m2 | 13454 | 11 | 96.7% (93.8-98.2) | 98.2% (96.6-99) | 98.2% (96.7-99) |
| **1951-1970 (50-69)** | Unvaccinated | 748546 | 2009 | [ref] | [ref] | [ref] |
| **1951-1970 (50-69)** | Primary series partly | 265532 | 145 | 75.6% (70.8-79.6) | 82.6% (79.1-85.6) | 84.2% (81-86.9) |
| **1951-1970 (50-69)** | Primary series m1 | 119387 | <10 | 96.5% (92.6-98.4) | 96.8% (93.2-98.5) | 97.2% (94.1-98.7) |
| **1951-1970 (50-69)** | Primary series m2 | 118579 | <10 | 97.1% (94.1-98.6) | 97.4% (94.8-98.7) | 97.7% (95.2-98.9) |
| **1951-1970 (50-69)** | Primary series m3 | 116455 | 20 | 93.2% (89.2-95.7) | 94.2% (90.7-96.3) | 94.8% (91.7-96.7) |
| **1951-1970 (50-69)** | Primary series m4 | 115809 | 36 | 92.4% (89.3-94.6) | 92.2% (89-94.5) | 93.1% (90.2-95.1) |
| **1951-1970 (50-69)** | Primary series m5 | 113602 | 81 | 91.2% (88.8-93) | 91.6% (89.3-93.4) | 92.5% (90.4-94.1) |

| | | | | | | |
|---|---|---|---|---|---|---|
| **1951-1970 (50-69)** | Primary series m6 | 93957 | 123 | 85.3% (82-88) | 89.4% (87-91.4) | 90.6% (88.5-92.3) |
| **1951-1970 (50-69)** | Primary series m7 | 36851 | 62 | 75.4% (67.8-81.2) | 84.9% (80.1-88.5) | 88.1% (84.4-90.9) |
| **1951-1970 (50-69)** | Primary series m8 | 9481 | 44 | 9.5% (-23.5-33.7) | 31.5% (6.3-50) | 63.9% (50.3-73.7) |
| **1951-1970 (50-69)** | Primary series m9 | 5119 | 38 | 4.4% (-33.5-31.5) | -17.8% (-64.4-15.6) | 23.7% (-6.9-45.6) |
| **1951-1970 (50-69)** | Primary series m10 | 3725 | <10 | 74.5% (50.6-86.8) | 60.7% (23.8-79.7) | 65.5% (33.2-82.2) |
| **1951-1970 (50-69)** | Booster m1 | 76888 | 17 | 95.7% (93-97.4) | 96.8% (94.7-98.1) | 96.9% (94.9-98.1) |
| **1951-1970 (50-69)** | Booster m2 | 7503 | <10 | 91.5% (72.9-97.3) | 93.5% (79.2-98) | 93.6% (79.5-98) |
| **1971-2009 (12-49)** | Unvaccinated | 1992705 | 154 | [ref] | [ref] | [ref] |
| **1971-2009 (12-49)** | Primary series partly | 366041 | <10 | 74.6% (47.4-87.8) | 75.9% (49.6-88.5) | 77.7% (53.5-89.3) |
| **1971-2009 (12-49)** | Primary series m1 | 164614 | <10 | 94.4% (59.4-99.2) | 95.8% (68.9-99.4) | 95.8% (69.1-99.4) |
| **1971-2009 (12-49)** | Primary series m2 | 157578 | <10 | 94.3% (58.5-99.2) | 95.9% (69.8-99.4) | 96.3% (72.8-99.5) |
| **1971-2009 (12-49)** | Primary series m3 | 153765 | <10 | 89% (54.9-97.3) | 89.8% (57.2-97.6) | 91.2% (63.4-97.9) |
| **1971-2009 (12-49)** | Primary series m4 | 146518 | <10 | 87.7% (65.9-95.6) | 89.5% (70.5-96.3) | 89.8% (71.6-96.4) |
| **1971-2009 (12-49)** | Primary series m5 | 134024 | <10 | 93.5% (73.2-98.4) | 96.2% (84.3-99.1) | 96.5% (85.6-99.2) |

Figure S1. Four-week moving average of number of COVID-19 deaths per week (A) and incidence per 100.000 person-days (B), per stratum of long-term care use and birth cohort. Data points based on fewer than 10 deaths are not shown to prevent disclosure of groups or persons.

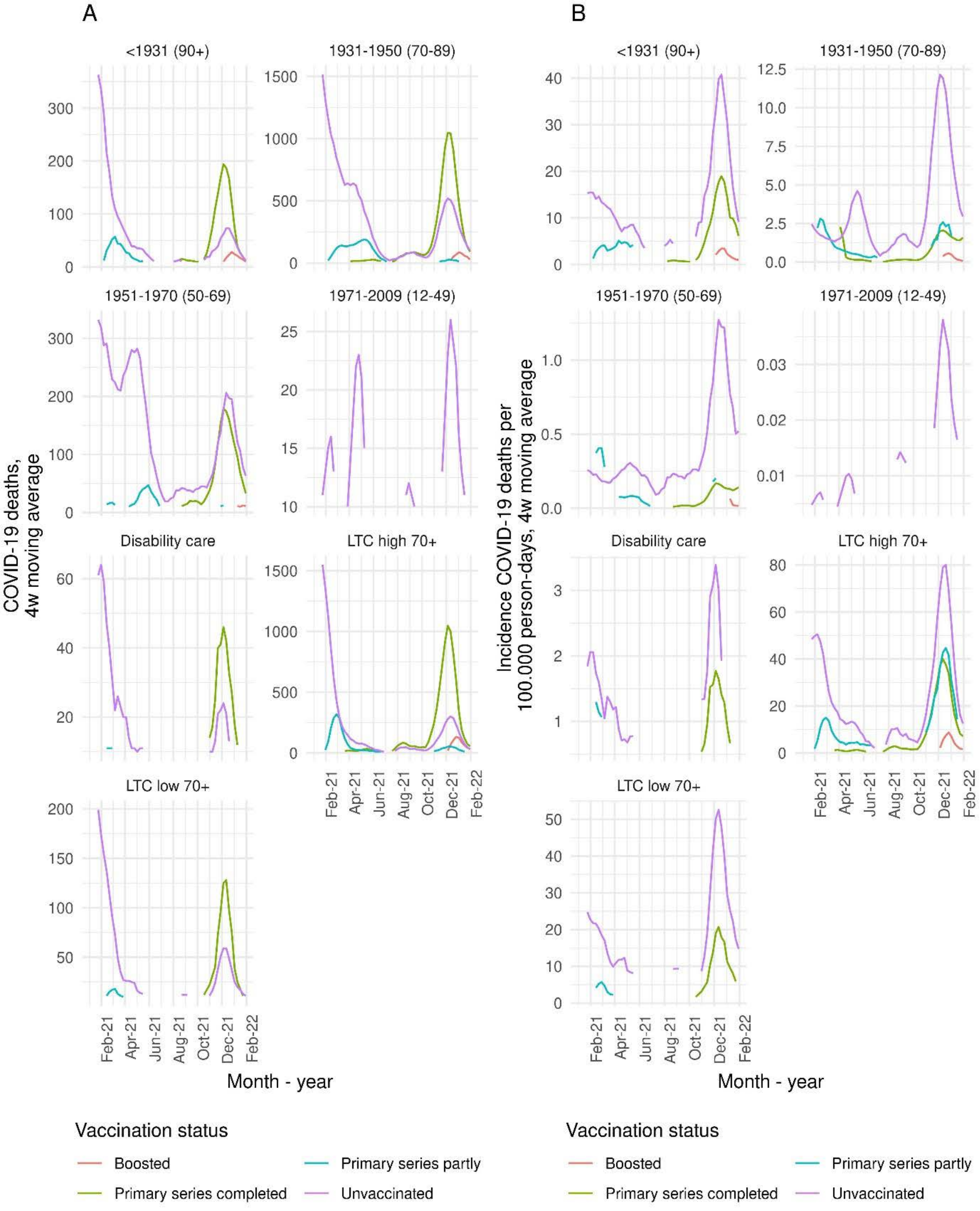

Figure S2. VE estimates of primary and booster vaccination against all-cause mortality, with 95% confidence interval, per stratum of long-term care use and birth cohort, adjusted for sex, year of birth, medical risk and country of origin. Underlying numbers can be found in Table S2 (as VE adjusted2).

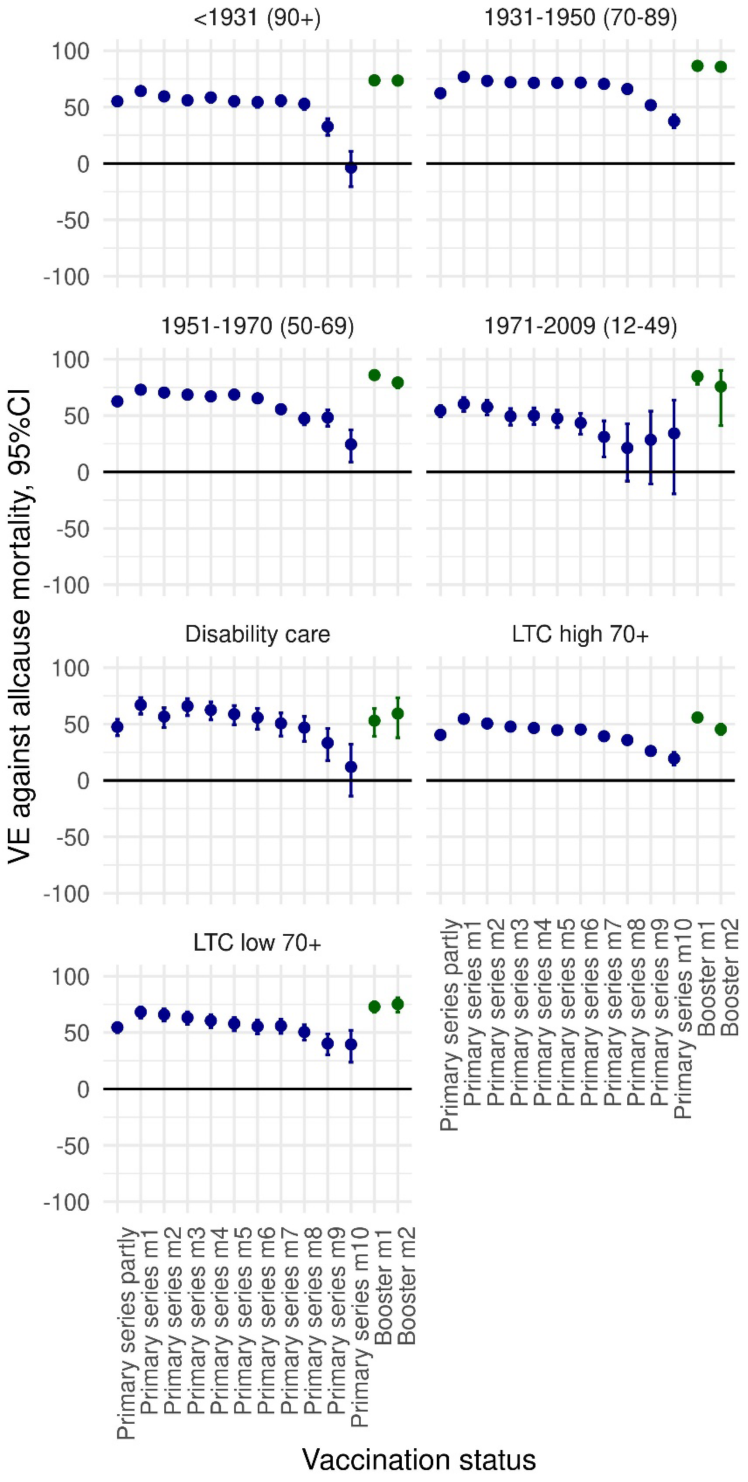

Figure S3. Four-week moving average of number of non-COVID-19 deaths per week (A) and incidence per 100.000 person-days (B), per stratum of long-term care use and birth cohort. Data points based on fewer than 10 deaths are not shown to prevent disclosure of groups or persons.

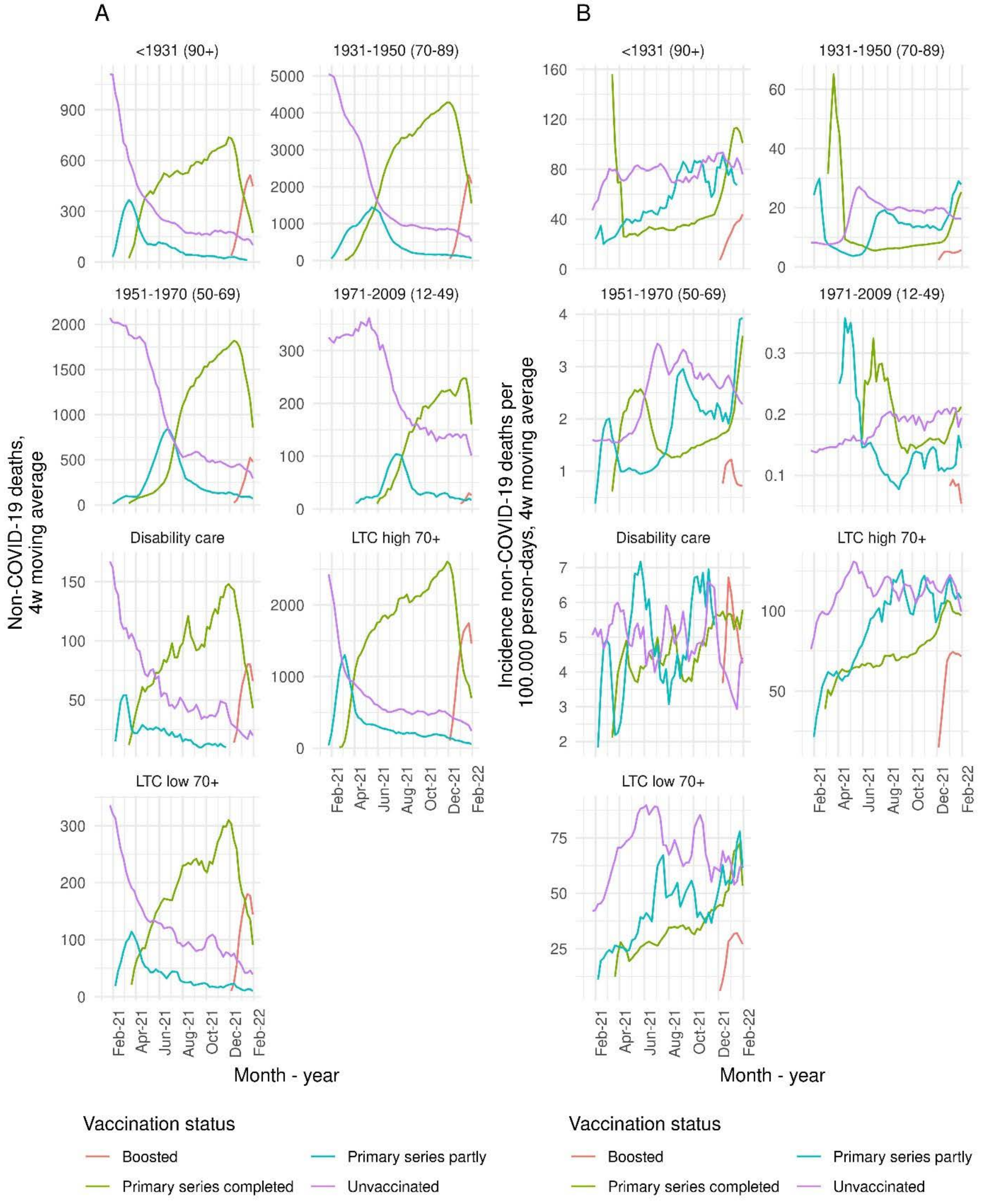

Table S2. VE estimates of primary and booster vaccination against all-cause mortality, with 95% confidence interval, per stratum of long-term care use and birth cohort, crude, adjusted for sex, year of birth and country of origin (adjusted1) and adjusted for sex, year of birth, country of origin and medical risk (adjusted2). Numbers below 10 are not shown to prevent disclosure of groups or persons.

| Stratum | Vaccination status | Person-time (x 1000 days) | Deaths | VE (95% CI) crude | VE (95% CI) adjusted1 | VE (95% CI) adjusted2 |
|---|---|---|---|---|---|---|
| **LTC high 70+** | Unvaccinated | 10778 | 13973 | [ref] | [ref] | [ref] |
| **LTC high 70+** | Primary series partly | 6368 | 4959 | 39.6% (37.4-41.7) | 40.5% (38.3-42.5) | 40.4% (38.3-42.5) |
| **LTC high 70+** | Primary series m1 | 3361 | 2001 | 53.9% (51.4-56.3) | 54.7% (52.2-57) | 54.6% (52.2-57) |
| **LTC high 70+** | Primary series m2 | 3291 | 2156 | 49.9% (47.2-52.5) | 50.5% (47.8-53.1) | 50.5% (47.8-53) |
| **LTC high 70+** | Primary series m3 | 3338 | 2296 | 47.3% (44.5-49.9) | 47.7% (44.9-50.4) | 47.7% (44.9-50.4) |
| **LTC high 70+** | Primary series m4 | 3311 | 2335 | 45.9% (43.1-48.6) | 46.5% (43.7-49.2) | 46.5% (43.7-49.2) |
| **LTC high 70+** | Primary series m5 | 3368 | 2558 | 43.8% (40.9-46.5) | 44.6% (41.7-47.3) | 44.6% (41.7-47.3) |
| **LTC high 70+** | Primary series m6 | 3310 | 2531 | 44.4% (41.5-47.1) | 45.1% (42.3-47.8) | 45.2% (42.3-47.8) |
| **LTC high 70+** | Primary series m7 | 3113 | 2700 | 37.4% (34.3-40.4) | 39.1% (36.1-42.1) | 39.2% (36.1-42.2) |
| **LTC high 70+** | Primary series m8 | 2779 | 2912 | 31.7% (28.3-35) | 35.8% (32.5-38.9) | 35.8% (32.5-38.9) |
| **LTC high 70+** | Primary series m9 | 1917 | 2670 | 20.2% (15.9-24.3) | 26.2% (22.2-30) | 26.1% (22.1-29.9) |
| **LTC high 70+** | Primary series m10 | 933 | 1285 | 12% (5.6-18) | 19.6% (13.8-25.1) | 19.4% (13.5-24.8) |
| **LTC high 70+** | Booster m1 | 2629 | 2069 | 52.5% (49.5-55.3) | 56% (53.2-58.6) | 55.8% (53-58.4) |
| **LTC high 70+** | Booster m2 | 1401 | 1179 | 40% (34.7-44.8) | 45.8% (41-50.1) | 45.4% (40.7-49.8) |
| **LTC low 70+** | Unvaccinated | 3159 | 2386 | [ref] | [ref] | [ref] |
| **LTC low 70+** | Primary series partly | 1556 | 550 | 53.5% (48.7-57.9) | 54.1% (49.4-58.4) | 54.6% (49.9-58.9) |
| **LTC low 70+** | Primary series m1 | 783 | 200 | 67.4% (61.9-72.1) | 68% (62.6-72.6) | 68.2% (62.9-72.8) |
| **LTC low 70+** | Primary series m2 | 767 | 216 | 65.3% (59.5-70.2) | 65.6% (59.9-70.5) | 65.9% (60.2-70.8) |
| **LTC low 70+** | Primary series m3 | 765 | 239 | 62.6% (56.6-67.7) | 62.8% (56.9-68) | 63.2% (57.3-68.3) |
| **LTC low 70+** | Primary series m4 | 763 | 258 | 59.8% (53.6-65.2) | 60.1% (53.9-65.4) | 60.5% (54.4-65.8) |
| **LTC low 70+** | Primary series m5 | 764 | 278 | 57% (50.6-62.6) | 57.4% (51-63) | 57.9% (51.6-63.4) |
| **LTC low 70+** | Primary series m6 | 744 | 289 | 54.5% (47.7-60.3) | 54.7% (48-60.6) | 55.4% (48.8-61.2) |
| **LTC low 70+** | Primary series m7 | 702 | 291 | 54.3% (47.5-60.3) | 55.2% (48.5-61) | 55.9% (49.3-61.7) |
| **LTC low 70+** | Primary series m8 | 596 | 331 | 46.2% (38.3-53.1) | 49.7% (42.3-56.1) | 50.5% (43.2-56.8) |

| | | | | | | |
|---|---|---|---|---|---|---|
| **LTC low 70+** | Primary series m9 | 324 | 251 | 32.1% (20.8-41.8) | 39.5% (29.4-48.1) | 40.3% (30.3-48.8) |
| **LTC low 70+** | Primary series m10 | 122 | 97 | 28.3% (9.7-43.1) | 38.6% (22.6-51.3) | 39.5% (23.7-52) |
| **LTC low 70+** | Booster m1 | 657 | 224 | 69.8% (64.1-74.6) | 72.8% (67.7-77.1) | 72.9% (67.9-77.2) |
| **LTC low 70+** | Booster m2 | 302 | 96 | 70.9% (62.5-77.5) | 75.2% (68-80.8) | 75.2% (68-80.8) |
| **Disability care** | Unvaccinated | 18128 | 1116 | [ref] | [ref] | [ref] |
| **Disability care** | Primary series partly | 5744 | 273 | 21.9% (10.5-31.8) | 47.5% (39.7-54.2) | 47.5% (39.7-54.2) |
| **Disability care** | Primary series m1 | 2948 | 94 | 47.5% (34.8-57.8) | 67.6% (59.6-74) | 67% (58.9-73.5) |
| **Disability care** | Primary series m2 | 2895 | 123 | 27.3% (11.6-40.2) | 57.2% (47.7-65) | 56.6% (46.9-64.4) |
| **Disability care** | Primary series m3 | 2861 | 99 | 41% (26.8-52.4) | 66.4% (58.1-73.1) | 65.9% (57.5-72.6) |
| **Disability care** | Primary series m4 | 2827 | 112 | 32.3% (16.7-45) | 62.9% (54.2-70) | 62.5% (53.7-69.6) |
| **Disability care** | Primary series m5 | 2770 | 121 | 20.1% (2-34.8) | 59.4% (50.1-67) | 58.8% (49.4-66.4) |
| **Disability care** | Primary series m6 | 2468 | 123 | 4.6% (-17.3-22.4) | 56.5% (46.5-64.7) | 55.7% (45.5-64) |
| **Disability care** | Primary series m7 | 2150 | 133 | -14.2% (-40.3-7) | 52% (41.2-60.9) | 50.6% (39.4-59.8) |
| **Disability care** | Primary series m8 | 1913 | 140 | -27% (-56.2--3.3) | 49.8% (38.4-59.1) | 46.8% (34.7-56.7) |
| **Disability care** | Primary series m9 | 1497 | 140 | -63.3% (-102--32.1) | 39.5% (25.4-51) | 33.3% (17.6-46.1) |
| **Disability care** | Primary series m10 | 794 | 91 | -142% (-212.7--87.3) | 23.7% (1.5-40.9) | 12% (-13.9-32.1) |
| **Disability care** | Booster m1 | 1918 | 108 | -26.7% (-64-2.1) | 57.1% (44.6-66.8) | 53% (39.2-63.7) |
| **Disability care** | Booster m2 | 580 | 32 | -29.5% (-97.9-15.3) | 64.4% (45.7-76.7) | 59.3% (37.9-73.4) |
| **<1931 (90+)** | Unvaccinated | 7375 | 5766 | [ref] | [ref] | [ref] |
| **<1931 (90+)** | Primary series partly | 3857 | 1368 | 55.8% (52.9-58.6) | 54.5% (51.4-57.3) | 55.1% (52.2-57.9) |
| **<1931 (90+)** | Primary series m1 | 2004 | 570 | 65.3% (61.8-68.5) | 63.7% (60-67) | 64.2% (60.6-67.5) |
| **<1931 (90+)** | Primary series m2 | 1936 | 625 | 60.7% (56.8-64.3) | 58.8% (54.6-62.5) | 59.5% (55.5-63.3) |
| **<1931 (90+)** | Primary series m3 | 1915 | 687 | 57.2% (52.9-61.1) | 55% (50.5-59) | 56% (51.6-59.9) |
| **<1931 (90+)** | Primary series m4 | 1853 | 631 | 59.6% (55.4-63.4) | 57.5% (53.1-61.5) | 58.6% (54.3-62.5) |
| **<1931 (90+)** | Primary series m5 | 1835 | 687 | 56.1% (51.6-60.1) | 53.9% (49.3-58.1) | 55.2% (50.7-59.3) |
| **<1931 (90+)** | Primary series m6 | 1732 | 690 | 55.3% (50.8-59.5) | 53% (48.2-57.4) | 54.4% (49.8-58.6) |
| **<1931 (90+)** | Primary series m7 | 1567 | 672 | 56.8% (52.2-60.9) | 54.2% (49.3-58.6) | 55.7% (51.1-60) |
| **<1931 (90+)** | Primary series m8 | 1413 | 751 | 53.9% (49-58.3) | 50.9% (45.7-55.6) | 52.8% (47.8-57.3) |
| **<1931 (90+)** | Primary series m9 | 671 | 600 | 33.4% (25.8-40.2) | 29.6% (21.6-36.9) | 32.7% (25-39.6) |

| | | | | | | |
|---|---|---|---|---|---|---|
| **<1931 (90+)** | Primary series m10 | 157 | 244 | -7.9% (-25.4-7.1) | -12.7% (-31-3) | -3.7% (-20.6-10.8) |
| **<1931 (90+)** | Booster m1 | 1462 | 536 | 74.1% (71.1-76.8) | 72.8% (69.6-75.7) | 73.7% (70.6-76.5) |
| **<1931 (90+)** | Booster m2 | 891 | 331 | 74.1% (70-77.6) | 72.7% (68.4-76.4) | 73.5% (69.4-77.1) |
| **1931-1950 (70-89)** | Unvaccinated | 250627 | 32578 | [ref] | [ref] | [ref] |
| **1931-1950 (70-89)** | Primary series partly | 108277 | 7316 | 61.6% (60.4-62.7) | 60.6% (59.5-61.7) | 62.3% (61.2-63.3) |
| **1931-1950 (70-89)** | Primary series m1 | 59638 | 3030 | 76% (75-77) | 75.6% (74.6-76.6) | 76.8% (75.8-77.7) |
| **1931-1950 (70-89)** | Primary series m2 | 59665 | 3713 | 71.7% (70.5-72.8) | 71.7% (70.5-72.7) | 73.2% (72.2-74.3) |
| **1931-1950 (70-89)** | Primary series m3 | 59546 | 3951 | 69.9% (68.7-71.1) | 70.2% (69-71.3) | 72% (70.9-73.1) |
| **1931-1950 (70-89)** | Primary series m4 | 58656 | 4074 | 68.2% (67-69.5) | 69.4% (68.2-70.5) | 71.5% (70.4-72.6) |
| **1931-1950 (70-89)** | Primary series m5 | 58535 | 4273 | 66.7% (65.4-68) | 69.1% (68-70.3) | 71.5% (70.4-72.6) |
| **1931-1950 (70-89)** | Primary series m6 | 57014 | 4590 | 63.9% (62.4-65.2) | 68.9% (67.7-70) | 71.7% (70.6-72.7) |
| **1931-1950 (70-89)** | Primary series m7 | 46666 | 4663 | 54.4% (52.5-56.2) | 67.3% (66-68.5) | 70.5% (69.3-71.6) |
| **1931-1950 (70-89)** | Primary series m8 | 20076 | 3577 | 20.6% (17.1-24) | 60.6% (59-62.3) | 66% (64.5-67.4) |
| **1931-1950 (70-89)** | Primary series m9 | 4768 | 1737 | -65.2% (-74.8--56.1) | 38.8% (35.4-42.2) | 51.7% (48.9-54.3) |
| **1931-1950 (70-89)** | Primary series m10 | 801 | 573 | -204.2% (-233.1--177.7) | 8.9% (0.3-16.7) | 37.5% (31.6-42.9) |
| **1931-1950 (70-89)** | Booster m1 | 48724 | 2464 | 78.1% (76.8-79.2) | 86.5% (85.8-87.2) | 86.5% (85.8-87.2) |
| **1931-1950 (70-89)** | Booster m2 | 13454 | 1057 | 67.5% (64.8-69.9) | 86.1% (85-87.1) | 85.7% (84.6-86.8) |
| **1951-1970 (50-69)** | Unvaccinated | 748546 | 16411 | [ref] | [ref] | [ref] |
| **1951-1970 (50-69)** | Primary series partly | 265532 | 3576 | 50% (47.9-52.1) | 59.3% (57.5-61) | 62.6% (60.9-64.1) |
| **1951-1970 (50-69)** | Primary series m1 | 119387 | 1369 | 62.7% (60.3-65) | 69.7% (67.7-71.6) | 72.9% (71.1-74.6) |
| **1951-1970 (50-69)** | Primary series m2 | 118579 | 1590 | 58% (55.4-60.4) | 66.4% (64.3-68.4) | 70.4% (68.6-72.1) |
| **1951-1970 (50-69)** | Primary series m3 | 116455 | 1718 | 52.9% (50-55.6) | 63.2% (60.9-65.3) | 68.5% (66.6-70.3) |
| **1951-1970 (50-69)** | Primary series m4 | 115809 | 1828 | 49.6% (46.6-52.4) | 60.2% (57.8-62.5) | 67% (65-68.8) |
| **1951-1970 (50-69)** | Primary series m5 | 113602 | 1826 | 51.1% (48.1-53.9) | 61.9% (59.6-64.1) | 68.6% (66.7-70.4) |
| **1951-1970 (50-69)** | Primary series m6 | 93957 | 1842 | 41% (37.4-44.4) | 56.9% (54.2-59.5) | 65.3% (63.2-67.3) |
| **1951-1970 (50-69)** | Primary series m7 | 36851 | 1176 | 5% (-1.8-11.4) | 37.8% (33.2-42) | 55.6% (52.4-58.6) |
| **1951-1970 (50-69)** | Primary series m8 | 9481 | 490 | -55.2% (-70.7--41.1) | -13.8% (-25.3--3.3) | 47.2% (41.8-52.1) |
| **1951-1970 (50-69)** | Primary series m9 | 5119 | 223 | -29.4% (-48.3--12.8) | -21.5% (-39.3--6) | 48.3% (40.6-55) |
| **1951-1970 (50-69)** | Primary series m10 | 3725 | 114 | 10.3% (-8.3-25.6) | 2.8% (-17.3-19.5) | 24.5% (8.8-37.4) |

| | | | | | | |
|---|---|---|---|---|---|---|
| **1951-1970 (50-69)** | Booster m1 | 76888 | 628 | 77.2% (75-79.3) | 84.8% (83.4-86.2) | 85.9% (84.6-87.2) |
| **1951-1970 (50-69)** | Booster m2 | 7503 | 107 | 65.5% (57.7-71.8) | 78.1% (73.2-82.2) | 79.3% (74.6-83.1) |
| **1971-2009 (12-49)** | Unvaccinated | 1992705 | 3424 | [ref] | [ref] | [ref] |
| **1971-2009 (12-49)** | Primary series partly | 366041 | 447 | 38% (31-44.3) | 50.7% (45-55.7) | 54.1% (49-58.8) |
| **1971-2009 (12-49)** | Primary series m1 | 164614 | 194 | 41% (31.2-49.5) | 55.8% (48.3-62.3) | 60.3% (53.7-66.1) |
| **1971-2009 (12-49)** | Primary series m2 | 157578 | 203 | 34.4% (23.5-43.7) | 51.4% (43.1-58.4) | 57.6% (50.6-63.6) |
| **1971-2009 (12-49)** | Primary series m3 | 153765 | 238 | 19.1% (6.5-30.1) | 40.3% (30.8-48.5) | 49.4% (41.5-56.2) |
| **1971-2009 (12-49)** | Primary series m4 | 146518 | 237 | 15.5% (2.1-27.1) | 40.9% (31.4-49.2) | 50% (42.1-56.8) |
| **1971-2009 (12-49)** | Primary series m5 | 134024 | 248 | 1.8% (-13.8-15.3) | 36.5% (26.3-45.4) | 47.6% (39.4-54.7) |
| **1971-2009 (12-49)** | Primary series m6 | 68084 | 190 | -48.4% (-75.1--25.8) | 20.8% (6.3-33) | 43.5% (33.4-52) |
| **1971-2009 (12-49)** | Primary series m7 | 18495 | 83 | -134.2% (-194.1--86.5) | -37.4% (-72.9--9.1) | 31.1% (13.3-45.3) |
| **1971-2009 (12-49)** | Primary series m8 | 7506 | 41 | -188.7% (-295.3--110.8) | -94.4% (-166.4--41.8) | 21.3% (-8.1-42.7) |
| **1971-2009 (12-49)** | Primary series m9 | 4890 | 21 | -123.7% (-245.9--44.7) | -51.4% (-134.3-2.2) | 28.5% (-10.6-53.9) |
| **1971-2009 (12-49)** | Primary series m10 | 4509 | 11 | -29.4% (-135.1-28.8) | 11% (-61.8-51.1) | 34.2% (-19.5-63.8) |
| **1971-2009 (12-49)** | Booster m1 | 48508 | 34 | 62% (45.7-73.4) | 81.3% (73.2-86.9) | 84.6% (77.9-89.2) |
| **1971-2009 (12-49)** | Booster m2 | 5199 | <10 | 48.9% (-24-78.9) | 70.4% (28.2-87.8) | 75.7% (41-90) |

Table S3. Hazard ratio (HR) estimates of the risk of non-COVID-19 mortality after a first dose of COVID-19 mRNA vaccine, compared to unvaccinated, with 95% confidence interval, per stratum of long-term care use and birth cohort, crude, adjusted for sex, year of birth and country of origin (adjusted1) and adjusted for sex, year of birth, country of origin and medical risk (adjusted2). Numbers below 10 are not shown to prevent disclosure of groups or persons.

| Stratum | Risk period | Person-time (x 1000 days) | Non-COVID-19 deaths | HR (95% CI) crude | HR (95% CI) adjusted1 | HR (95% CI) adjusted2 |
|---|---|---:|---|---|---|---|
| **LTC high 70+** | reference | 10772 | 10361 | [ref] | [ref] | [ref] |
| **LTC high 70+** | week 1 | 761 | 212 | 0.3 (0.2-0.3) | 0.3 (0.2-0.3) | 0.3 (0.2-0.3) |
| **LTC high 70+** | week 2 | 761 | 394 | 0.5 (0.4-0.5) | 0.5 (0.4-0.5) | 0.5 (0.4-0.5) |
| **LTC high 70+** | week 3 | 760 | 455 | 0.6 (0.5-0.6) | 0.6 (0.5-0.6) | 0.6 (0.5-0.6) |
| **LTC high 70+** | week 4 | 720 | 420 | 0.6 (0.5-0.6) | 0.5 (0.5-0.6) | 0.5 (0.5-0.6) |
| **LTC high 70+** | week 5 | 266 | 258 | 0.9 (0.8-1) | 0.9 (0.8-1) | 0.9 (0.8-1) |
| **LTC low 70+** | reference | 3157 | 1791 | [ref] | [ref] | [ref] |
| **LTC low 70+** | week 1 | 179 | 22 | 0.2 (0.1-0.3) | 0.2 (0.1-0.3) | 0.2 (0.1-0.3) |
| **LTC low 70+** | week 2 | 178 | 32 | 0.3 (0.2-0.4) | 0.3 (0.2-0.4) | 0.3 (0.2-0.4) |
| **LTC low 70+** | week 3 | 177 | 33 | 0.3 (0.2-0.4) | 0.3 (0.2-0.4) | 0.3 (0.2-0.4) |
| **LTC low 70+** | week 4 | 171 | 51 | 0.5 (0.4-0.6) | 0.5 (0.4-0.6) | 0.5 (0.3-0.6) |
| **LTC low 70+** | week 5 | 110 | 33 | 0.4 (0.3-0.6) | 0.4 (0.3-0.6) | 0.4 (0.3-0.6) |
| **Disability care** | reference | 18126 | 869 | [ref] | [ref] | [ref] |
| **Disability care** | week 1 | 666 | <10 | 0.2 (0.1-0.4) | 0.1 (0.1-0.3) | 0.1 (0.1-0.3) |
| **Disability care** | week 2 | 660 | 15 | 0.4 (0.3-0.7) | 0.3 (0.2-0.5) | 0.3 (0.2-0.5) |
| **Disability care** | week 3 | 656 | 27 | 0.8 (0.5-1.2) | 0.5 (0.4-0.8) | 0.6 (0.4-0.8) |
| **Disability care** | week 4 | 572 | 18 | 0.6 (0.4-1) | 0.4 (0.3-0.7) | 0.4 (0.3-0.7) |
| **Disability care** | week 5 | 285 | 13 | 1 (0.6-1.7) | 0.7 (0.4-1.1) | 0.6 (0.4-1.1) |
| **<1931 (90+)** | reference | 7374 | 4718 | [ref] | [ref] | [ref] |
| **<1931 (90+)** | week 1 | 443 | 49 | 0.2 (0.1-0.2) | 0.2 (0.1-0.2) | 0.2 (0.1-0.2) |
| **<1931 (90+)** | week 2 | 441 | 98 | 0.3 (0.3-0.4) | 0.3 (0.3-0.4) | 0.3 (0.3-0.4) |
| **<1931 (90+)** | week 3 | 438 | 96 | 0.3 (0.2-0.4) | 0.3 (0.2-0.4) | 0.3 (0.2-0.4) |
| **<1931 (90+)** | week 4 | 433 | 109 | 0.4 (0.3-0.4) | 0.4 (0.3-0.4) | 0.4 (0.3-0.4) |
| **<1931 (90+)** | week 5 | 416 | 114 | 0.4 (0.3-0.5) | 0.4 (0.3-0.5) | 0.4 (0.3-0.5) |

| | | | | | | |
|---|---|---|---|---|---|---|
| **1931-1950 (70-89)** | reference | 250621 | 25637 | [ref] | [ref] | [ref] |
| **1931-1950 (70-89)** | week 1 | 13833 | 312 | 0.2 (0.2-0.2) | 0.2 (0.2-0.2) | 0.2 (0.2-0.2) |
| **1931-1950 (70-89)** | week 2 | 13821 | 454 | 0.3 (0.2-0.3) | 0.2 (0.2-0.3) | 0.2 (0.2-0.2) |
| **1931-1950 (70-89)** | week 3 | 13808 | 564 | 0.3 (0.3-0.3) | 0.3 (0.2-0.3) | 0.3 (0.2-0.3) |
| **1931-1950 (70-89)** | week 4 | 13722 | 599 | 0.3 (0.3-0.3) | 0.3 (0.2-0.3) | 0.3 (0.2-0.3) |
| **1931-1950 (70-89)** | week 5 | 13427 | 652 | 0.3 (0.3-0.3) | 0.3 (0.3-0.3) | 0.3 (0.3-0.3) |
| **1951-1970 (50-69)** | reference | 748541 | 13458 | [ref] | [ref] | [ref] |
| **1951-1970 (50-69)** | week 1 | 19105 | 94 | 0.2 (0.2-0.3) | 0.2 (0.2-0.2) | 0.2 (0.1-0.2) |
| **1951-1970 (50-69)** | week 2 | 19096 | 131 | 0.3 (0.3-0.4) | 0.3 (0.2-0.3) | 0.2 (0.2-0.3) |
| **1951-1970 (50-69)** | week 3 | 19085 | 163 | 0.4 (0.3-0.4) | 0.3 (0.2-0.3) | 0.2 (0.2-0.3) |
| **1951-1970 (50-69)** | week 4 | 18206 | 165 | 0.4 (0.3-0.4) | 0.3 (0.2-0.3) | 0.2 (0.2-0.3) |
| **1951-1970 (50-69)** | week 5 | 17241 | 181 | 0.4 (0.4-0.5) | 0.3 (0.3-0.4) | 0.3 (0.2-0.3) |
| **1971-2009 (12-49)** | reference | 1992704 | 2967 | [ref] | [ref] | [ref] |
| **1971-2009 (12-49)** | week 1 | 36934 | 28 | 0.5 (0.3-0.7) | 0.4 (0.3-0.6) | 0.3 (0.2-0.5) |
| **1971-2009 (12-49)** | week 2 | 36893 | 31 | 0.5 (0.4-0.8) | 0.4 (0.3-0.6) | 0.4 (0.2-0.5) |
| **1971-2009 (12-49)** | week 3 | 36831 | 31 | 0.5 (0.4-0.8) | 0.4 (0.3-0.6) | 0.3 (0.2-0.5) |

| | | | | | | |
|---|---|---|---|---|---|---|
| **1971-2009 (12-49)** | week 4 | 33513 | 38 | 0.7 (0.5-1) | 0.5 (0.3-0.7) | 0.4 (0.3-0.6) |
| **1971-2009 (12-49)** | week 5 | 30261 | 41 | 0.9 (0.6-1.2) | 0.5 (0.4-0.7) | 0.5 (0.3-0.7) |

Table S4. Hazard ratio (HR) estimates of the risk of non-COVID-19 mortality after a first dose of COVID-19 vector vaccine, compared to unvaccinated, with 95% confidence interval, per stratum of long-term care use and birth cohort, crude, adjusted for sex, year of birth and country of origin (adjusted1) and adjusted for sex, year of birth, country of origin and medical risk (adjusted2). Numbers below 10 are not shown to prevent disclosure of groups or persons.

| **Stratum** | **Risk period** | **Person-time (x 1000 days)** | **Non-COVID-19 deaths** | **HR (95% CI) crude** | **HR (95% CI) adjusted1** | **HR (95% CI) adjusted2** |
|---|---|---|---|---|---|---|
| **LTC high 70+** | reference | 10772 | 10361 | [ref] | [ref] | [ref] |
| **LTC high 70+** | week 1 | 18 | <10 | 0.3 (0.1-0.6) | 0.3 (0.1-0.6) | 0.3 (0.1-0.6) |
| **LTC high 70+** | week 2 | 18 | 13 | 0.6 (0.3-1.0) | 0.6 (0.3-1.0) | 0.6 (0.3-1.0) |
| **LTC high 70+** | week 3 | 18 | 10 | 0.5 (0.2-0.9) | 0.4 (0.2-0.8) | 0.4 (0.2-0.8) |
| **LTC high 70+** | week 4 | 18 | 15 | 0.7 (0.4-1.2) | 0.6 (0.4-1.1) | 0.6 (0.4-1.1) |
| **LTC high 70+** | week 5 | 18 | 14 | 0.7 (0.4-1.1) | 0.6 (0.4-1.1) | 0.6 (0.4-1.1) |
| **LTC high 70+** | week 6 | 17 | 15 | 0.8 (0.5-1.3) | 0.7 (0.4-1.2) | 0.7 (0.4-1.2) |
| **LTC high 70+** | week 7 | 16 | 18 | 1 (0.6-1.6) | 0.9 (0.6-1.5) | 0.9 (0.6-1.5) |
| **LTC high 70+** | week 8 | 15 | 12 | 0.7 (0.4-1.3) | 0.7 (0.4-1.2) | 0.7 (0.4-1.2) |
| **LTC low 70+** | reference | 3157 | 1791 | [ref] | [ref] | [ref] |
| **LTC low 70+** | week 1 | 7 | <10 | 0.2 (0-1.3) | 0.2 (0-1.2) | 0.2 (0-1.2) |
| **LTC low 70+** | week 2 | 7 | <10 | 0.7 (0.3-1.9) | 0.7 (0.2-1.7) | 0.6 (0.2-1.7) |
| **LTC low 70+** | week 3 | 7 | <10 | 0.2 (0-1.3) | 0.2 (0-1.2) | 0.2 (0-1.1) |
| **LTC low 70+** | week 4 | 7 | <10 | 0.3 (0.1-1.4) | 0.3 (0.1-1.3) | 0.3 (0.1-1.2) |
| **LTC low 70+** | week 5 | 7 | <10 | 0.2 (0-1.2) | 0.2 (0-1.1) | 0.2 (0-1.1) |
| **LTC low 70+** | week 6 | 7 | <10 | 0.5 (0.2-1.6) | 0.5 (0.2-1.5) | 0.5 (0.2-1.5) |
| **LTC low 70+** | week 7 | 6 | <10 | 0.4 (0.1-1.6) | 0.4 (0.1-1.4) | 0.4 (0.1-1.4) |
| **LTC low 70+** | week 8 | 6 | <10 | 0.4 (0.1-1.7) | 0.4 (0.1-1.6) | 0.4 (0.1-1.5) |
| **Disability care** | reference | 18126 | 869 | [ref] | [ref] | [ref] |

| | | | | | | |
|---|---|---|---|---|---|---|
| **Disability care** | week 1 | 35 | <10 | 1.7 (0.5-5.2) | 1.1 (0.4-3.4) | 1 (0.3-3.2) |
| **Disability care** | week 2 | 35 | <10 | 0.6 (0.1-4.1) | 0.4 (0.1-2.6) | 0.3 (0-2.4) |
| **Disability care** | week 3 | 35 | <10 | <0.01 (--) | <0.01 (--) | <0.01 (--) |
| **Disability care** | week 4 | 35 | <10 | 1.8 (0.6-5.6) | 1.1 (0.3-3.3) | 1 (0.3-3.0) |
| **Disability care** | week 5 | 34 | <10 | 0.6 (0.1-4.0) | 0.3 (0-2.5) | 0.3 (0-2.3) |
| **Disability care** | week 6 | 34 | <10 | 0.6 (0.1-4.3) | 0.4 (0.1-2.7) | 0.3 (0-2.4) |
| **Disability care** | week 7 | 33 | <10 | 0.6 (0.1-4.3) | 0.4 (0.1-2.7) | 0.3 (0-2.4) |
| **Disability care** | week 8 | 33 | <10 | 0.7 (0.1-4.8) | 0.4 (0.1-3) | 0.4 (0.1-2.7) |
| **<1931 (90+)** | reference | 7374 | 4718 | [ref] | [ref] | [ref] |
| **<1931 (90+)** | week 1 | 42 | 13 | 0.4 (0.2-0.7) | 0.4 (0.2-0.7) | 0.4 (0.2-0.7) |
| **<1931 (90+)** | week 2 | 42 | <10 | 0.2 (0.1-0.5) | 0.2 (0.1-0.5) | 0.2 (0.1-0.5) |
| **<1931 (90+)** | week 3 | 41 | 12 | 0.4 (0.2-0.7) | 0.4 (0.2-0.6) | 0.4 (0.2-0.6) |
| **<1931 (90+)** | week 4 | 41 | <10 | 0.3 (0.1-0.5) | 0.3 (0.1-0.5) | 0.3 (0.1-0.5) |
| **<1931 (90+)** | week 5 | 39 | 17 | 0.6 (0.3-0.9) | 0.5 (0.3-0.9) | 0.5 (0.3-0.9) |
| **<1931 (90+)** | week 6 | 38 | 11 | 0.4 (0.2-0.7) | 0.4 (0.2-0.7) | 0.4 (0.2-0.7) |
| **<1931 (90+)** | week 7 | 35 | 17 | 0.6 (0.4-1.0) | 0.6 (0.4-1.0) | 0.6 (0.4-1.0) |
| **<1931 (90+)** | week 8 | 32 | 15 | 0.6 (0.4-1.0) | 0.6 (0.3-1.0) | 0.6 (0.3-1.0) |
| **1931-1950 (70-89)** | reference | 250621 | 25637 | [ref] | [ref] | [ref] |
| **1931-1950 (70-89)** | week 1 | 225 | 27 | 0.7 (0.5-1.1) | 0.5 (0.3-0.7) | 0.4 (0.3-0.6) |
| **1931-1950 (70-89)** | week 2 | 222 | 41 | 1 (0.7-1.3) | 0.7 (0.5-0.9) | 0.6 (0.4-0.8) |
| **1931-1950 (70-89)** | week 3 | 220 | 43 | 0.9 (0.7-1.3) | 0.7 (0.5-0.9) | 0.6 (0.5-0.8) |
| **1931-1950 (70-89)** | week 4 | 217 | 74 | 1.5 (1.2-1.9) | 1.2 (0.9-1.5) | 1 (0.8-1.3) |
| **1931-1950 (70-89)** | week 5 | 212 | 40 | 0.8 (0.6-1.1) | 0.6 (0.5-0.9) | 0.6 (0.4-0.8) |

| | | | | | | |
|---|---|---|---|---|---|---|
| **1931-1950 (70-89)** | week 6 | 204 | 58 | 1.2 (1-1.6) | 1 (0.8-1.3) | 0.9 (0.7-1.2) |
| **1931-1950 (70-89)** | week 7 | 193 | 61 | 1.4 (1.1-1.8) | 1.2 (0.9-1.5) | 1 (0.8-1.3) |
| **1931-1950 (70-89)** | week 8 | 180 | 62 | 1.6 (1.2-2) | 1.3 (1-1.7) | 1.2 (0.9-1.5) |
| **1951-1970 (50-69)** | reference | 748541 | 13458 | [ref] | [ref] | [ref] |
| **1951-1970 (50-69)** | week 1 | 9301 | 27 | 0.2 (0.1-0.2) | 0.1 (0.1-0.2) | 0.1 (0.1-0.2) |
| **1951-1970 (50-69)** | week 2 | 9254 | 74 | 0.4 (0.3-0.5) | 0.3 (0.3-0.4) | 0.3 (0.3-0.4) |
| **1951-1970 (50-69)** | week 3 | 9221 | 84 | 0.5 (0.4-0.6) | 0.4 (0.3-0.5) | 0.4 (0.3-0.4) |
| **1951-1970 (50-69)** | week 4 | 9195 | 72 | 0.4 (0.3-0.5) | 0.3 (0.2-0.4) | 0.3 (0.2-0.4) |
| **1951-1970 (50-69)** | week 5 | 9104 | 81 | 0.4 (0.3-0.5) | 0.3 (0.2-0.4) | 0.3 (0.2-0.4) |
| **1951-1970 (50-69)** | week 6 | 8937 | 95 | 0.5 (0.4-0.6) | 0.3 (0.3-0.4) | 0.3 (0.3-0.4) |
| **1951-1970 (50-69)** | week 7 | 8647 | 96 | 0.5 (0.4-0.6) | 0.3 (0.3-0.4) | 0.3 (0.3-0.4) |
| **1951-1970 (50-69)** | week 8 | 8278 | 93 | 0.5 (0.4-0.6) | 0.3 (0.3-0.4) | 0.3 (0.2-0.4) |
| **1971-2009 (12-49)** | reference | 1992704 | 2967 | [ref] | [ref] | [ref] |
| **1971-2009 (12-49)** | week 1 | 4373 | <10 | 0.6 (0.2-1.6) | 0.5 (0.2-1.3) | 0.5 (0.2-1.3) |
| **1971-2009 (12-49)** | week 2 | 4366 | <10 | 0.4 (0.1-1.3) | 0.4 (0.1-1.1) | 0.4 (0.1-1.1) |
| **1971-2009 (12-49)** | week 3 | 4363 | <10 | 0.3 (0.1-1.2) | 0.2 (0.1-1) | 0.2 (0.1-0.9) |

| | | | | | | |
|---|---|---|---|---|---|---|
| **1971-2009 (12-49)** | week 4 | 4358 | <10 | 0.6 (0.2-1.5) | 0.5 (0.2-1.2) | 0.5 (0.2-1.2) |
| **1971-2009 (12-49)** | week 5 | 4339 | <10 | 1.2 (0.6-2.4) | 0.9 (0.5-1.9) | 0.9 (0.5-1.9) |
| **1971-2009 (12-49)** | week 6 | 4310 | <10 | 0.1 (0-1.0) | 0.1 (0-0.8) | 0.1 (0-0.8) |
| **1971-2009 (12-49)** | week 7 | 4281 | <10 | 0.9 (0.4-2) | 0.7 (0.3-1.5) | 0.7 (0.3-1.5) |
| **1971-2009 (12-49)** | week 8 | 4249 | <10 | 0.6 (0.2-1.6) | 0.5 (0.2-1.3) | 0.5 (0.2-1.2) |

Table S5. Hazard ratio (HR) estimates of the risk of non-COVID-19 mortality after a second dose of COVID-19 vaccine, compared to person-time at least 2 weeks post-first dose, with 95% confidence interval, per stratum of long-term care use and birth cohort, crude, adjusted for sex, year of birth and country of origin (adjusted1) and adjusted for sex, year of birth, country of origin and medical risk (adjusted2). Numbers below 10 are not shown to prevent disclosure of groups or persons.

| Stratum | Risk period | Person-time (x 1000 days) | Non-COVID-19 deaths | HR (95% CI) crude | HR (95% CI) adjusted1 | HR (95% CI) adjusted2 |
|---|---|---|---|---|---|---|
| **LTC high 70+** | reference | 3327 | 3086 | [ref] | [ref] | [ref] |
| **LTC high 70+** | week 1 | 749 | 214 | 0.3 (0.2-0.3) | 0.3 (0.3-0.3) | 0.3 (0.3-0.3) |
| **LTC high 70+** | week 2 | 750 | 373 | 0.5 (0.4-0.5) | 0.5 (0.4-0.5) | 0.5 (0.4-0.5) |
| **LTC high 70+** | week 3 | 751 | 396 | 0.5 (0.5-0.6) | 0.6 (0.5-0.6) | 0.6 (0.5-0.6) |
| **LTC high 70+** | week 4 | 751 | 436 | 0.6 (0.5-0.7) | 0.6 (0.5-0.7) | 0.6 (0.5-0.7) |
| **LTC high 70+** | week 5 | 749 | 450 | 0.6 (0.5-0.7) | 0.6 (0.6-0.7) | 0.6 (0.6-0.7) |
| **LTC high 70+** | week 6 | 748 | 455 | 0.6 (0.6-0.7) | 0.6 (0.6-0.7) | 0.6 (0.6-0.7) |
| **LTC high 70+** | week 7 | 748 | 463 | 0.7 (0.6-0.7) | 0.7 (0.6-0.7) | 0.7 (0.6-0.7) |
| **LTC high 70+** | week 8 | 4451 | 2917 | 0.7 (0.7-0.7) | 0.7 (0.7-0.7) | 0.7 (0.7-0.7) |
| **LTC low 70+** | reference | 858 | 359 | [ref] | [ref] | [ref] |
| **LTC low 70+** | week 1 | 176 | 21 | 0.3 (0.2-0.5) | 0.3 (0.2-0.5) | 0.3 (0.2-0.5) |
| **LTC low 70+** | week 2 | 175 | 44 | 0.7 (0.5-0.9) | 0.7 (0.5-0.9) | 0.7 (0.5-0.9) |
| **LTC low 70+** | week 3 | 175 | 36 | 0.5 (0.4-0.8) | 0.5 (0.4-0.8) | 0.5 (0.4-0.8) |
| **LTC low 70+** | week 4 | 174 | 39 | 0.6 (0.4-0.8) | 0.6 (0.4-0.8) | 0.6 (0.4-0.8) |
| **LTC low 70+** | week 5 | 173 | 39 | 0.6 (0.4-0.8) | 0.6 (0.4-0.8) | 0.6 (0.4-0.8) |
| **LTC low 70+** | week 6 | 172 | 48 | 0.7 (0.5-0.9) | 0.7 (0.5-0.9) | 0.7 (0.5-1.0) |
| **LTC low 70+** | week 7 | 172 | 44 | 0.6 (0.4-0.9) | 0.6 (0.4-0.9) | 0.6 (0.4-0.9) |
| **LTC low 70+** | week 8 | 1020 | 297 | 0.7 (0.6-0.8) | 0.6 (0.5-0.8) | 0.6 (0.5-0.8) |
| **Disability care** | reference | 3216 | 181 | [ref] | [ref] | [ref] |
| **Disability care** | week 1 | 658 | 10 | 0.2 (0.1-0.4) | 0.3 (0.1-0.5) | 0.3 (0.1-0.5) |
| **Disability care** | week 2 | 655 | 22 | 0.5 (0.3-0.7) | 0.5 (0.3-0.8) | 0.5 (0.3-0.8) |
| **Disability care** | week 3 | 653 | 15 | 0.4 (0.2-0.6) | 0.4 (0.2-0.7) | 0.4 (0.2-0.7) |
| **Disability care** | week 4 | 650 | 15 | 0.4 (0.2-0.6) | 0.4 (0.2-0.7) | 0.4 (0.2-0.7) |

| | | | | | | |
|---|---|---|---|---|---|---|
| **Disability care** | week 5 | 644 | 25 | 0.7 (0.4-1.0) | 0.7 (0.4-1.0) | 0.7 (0.4-1.0) |
| **Disability care** | week 6 | 642 | 24 | 0.6 (0.4-0.9) | 0.6 (0.4-0.9) | 0.6 (0.4-0.9) |
| **Disability care** | week 7 | 640 | 19 | 0.5 (0.3-0.8) | 0.5 (0.3-0.8) | 0.5 (0.3-0.8) |
| **Disability care** | week 8 | 3805 | 153 | 0.7 (0.5-0.9) | 0.6 (0.5-0.8) | 0.6 (0.5-0.8) |
| **<1931 (90+)** | reference | 1959 | 872 | [ref] | [ref] | [ref] |
| **<1931 (90+)** | week 1 | 461 | 49 | 0.2 (0.2-0.3) | 0.2 (0.2-0.3) | 0.2 (0.2-0.3) |
| **<1931 (90+)** | week 2 | 459 | 111 | 0.4 (0.4-0.6) | 0.5 (0.4-0.6) | 0.5 (0.4-0.6) |
| **<1931 (90+)** | week 3 | 457 | 102 | 0.4 (0.3-0.5) | 0.4 (0.3-0.5) | 0.4 (0.3-0.5) |
| **<1931 (90+)** | week 4 | 454 | 121 | 0.5 (0.4-0.6) | 0.5 (0.4-0.6) | 0.5 (0.4-0.6) |
| **<1931 (90+)** | week 5 | 452 | 129 | 0.5 (0.4-0.6) | 0.5 (0.4-0.6) | 0.5 (0.4-0.6) |
| **<1931 (90+)** | week 6 | 449 | 128 | 0.5 (0.4-0.6) | 0.5 (0.4-0.6) | 0.5 (0.4-0.6) |
| **<1931 (90+)** | week 7 | 447 | 123 | 0.5 (0.4-0.6) | 0.5 (0.4-0.6) | 0.5 (0.4-0.6) |
| **<1931 (90+)** | week 8 | 2696 | 888 | 0.6 (0.5-0.7) | 0.6 (0.6-0.7) | 0.6 (0.6-0.7) |
| **1931-1950 (70-89)** | reference | 53088 | 4805 | [ref] | [ref] | [ref] |
| **1931-1950 (70-89)** | week 1 | 13728 | 327 | 0.3 (0.2-0.3) | 0.3 (0.2-0.3) | 0.3 (0.2-0.3) |
| **1931-1950 (70-89)** | week 2 | 13715 | 470 | 0.4 (0.3-0.4) | 0.4 (0.3-0.4) | 0.4 (0.3-0.4) |
| **1931-1950 (70-89)** | week 3 | 13702 | 558 | 0.5 (0.4-0.5) | 0.4 (0.4-0.4) | 0.4 (0.4-0.4) |
| **1931-1950 (70-89)** | week 4 | 13687 | 623 | 0.5 (0.5-0.6) | 0.4 (0.4-0.4) | 0.4 (0.4-0.5) |
| **1931-1950 (70-89)** | week 5 | 13670 | 692 | 0.6 (0.6-0.7) | 0.4 (0.4-0.5) | 0.4 (0.4-0.5) |
| **1931-1950 (70-89)** | week 6 | 13650 | 719 | 0.7 (0.6-0.8) | 0.4 (0.4-0.5) | 0.4 (0.4-0.5) |
| **1931-1950 (70-89)** | week 7 | 13627 | 818 | 0.8 (0.8-0.9) | 0.5 (0.4-0.5) | 0.5 (0.4-0.5) |
| **1931-1950 (70-89)** | week 8 | 82892 | 4907 | 0.8 (0.8-0.9) | 0.4 (0.3-0.4) | 0.4 (0.3-0.4) |

| | | | | | | |
|---|---|---|---|---|---|---|
| **1951-1970 (50-69)** | reference | 205705 | 2803 | [ref] | [ref] | [ref] |
| **1951-1970 (50-69)** | week 1 | 25384 | 137 | 0.4 (0.3-0.4) | 0.3 (0.3-0.4) | 0.3 (0.3-0.4) |
| **1951-1970 (50-69)** | week 2 | 25342 | 170 | 0.5 (0.4-0.5) | 0.4 (0.3-0.5) | 0.4 (0.3-0.4) |
| **1951-1970 (50-69)** | week 3 | 25309 | 261 | 0.7 (0.6-0.8) | 0.6 (0.5-0.7) | 0.6 (0.5-0.6) |
| **1951-1970 (50-69)** | week 4 | 25274 | 272 | 0.8 (0.7-0.9) | 0.6 (0.5-0.7) | 0.6 (0.5-0.6) |
| **1951-1970 (50-69)** | week 5 | 25233 | 279 | 0.8 (0.7-1.0) | 0.6 (0.5-0.7) | 0.6 (0.5-0.6) |
| **1951-1970 (50-69)** | week 6 | 25174 | 328 | 1.0 (0.9-1.2) | 0.7 (0.6-0.8) | 0.6 (0.6-0.7) |
| **1951-1970 (50-69)** | week 7 | 25119 | 321 | 1.1 (0.9-1.2) | 0.7 (0.6-0.8) | 0.6 (0.5-0.7) |
| **1951-1970 (50-69)** | week 8 | 152506 | 2043 | 1.2 (1.1-1.3) | 0.7 (0.7-0.8) | 0.6 (0.5-0.6) |
| **1971-2009 (12-49)** | reference | 292775 | 393 | [ref] | [ref] | [ref] |
| **1971-2009 (12-49)** | week 1 | 34400 | 13 | 0.3 (0.1-0.5) | 0.3 (0.2-0.5) | 0.3 (0.1-0.4) |
| **1971-2009 (12-49)** | week 2 | 34315 | 21 | 0.4 (0.3-0.7) | 0.4 (0.3-0.7) | 0.4 (0.3-0.6) |
| **1971-2009 (12-49)** | week 3 | 34220 | 30 | 0.7 (0.5-1.0) | 0.7 (0.5-1.0) | 0.6 (0.4-0.9) |
| **1971-2009 (12-49)** | week 4 | 34099 | 40 | 1.0 (0.7-1.4) | 1.0 (0.7-1.4) | 0.9 (0.6-1.2) |
| **1971-2009 (12-49)** | week 5 | 33971 | 43 | 1.1 (0.8-1.6) | 1.1 (0.8-1.6) | 0.9 (0.7-1.3) |
| **1971-2009 (12-49)** | week 6 | 33842 | 40 | 1.1 (0.8-1.5) | 1.1 (0.8-1.5) | 0.9 (0.6-1.2) |

| | | | | | | |
|---|---|---|---|---|---|---|
| **1971-2009 (12-49)** | week 7 | 33659 | 40 | 1.1 (0.8-1.6) | 1.1 (0.8-1.5) | 0.9 (0.6-1.2) |
| **1971-2009 (12-49)** | week 8 | 200360 | 254 | 1.3 (1.1-1.6) | 1.1 (0.9-1.4) | 0.9 (0.7-1.0) |

Table S6. Hazard ratio (HR) estimates with 95% confidence interval, for risk of non-COVID-19 mortality after a booster dose of COVID-19 vaccine, compared to person-time at least 3 months post-second dose, per stratum of long-term care use and birth cohort, crude, adjusted for sex, year of birth and country of origin (adjusted1) and adjusted for sex, year of birth, country of origin and medical risk (adjusted2). Numbers below 10 are not shown to prevent disclosure of groups or persons.

| Stratum | Risk period | Person-time (x 1000 days) | Non-COVID-19 deaths | HR (95% CI) crude | HR (95% CI) adjusted1 | HR (95% CI) adjusted2 |
|---|---|---|---|---|---|---|
| **LTC high 70+** | reference | 17187 | 14837 | [ref] | [ref] | [ref] |
| **LTC high 70+** | week 1 | 600 | 253 | 0.3 (0.3-0.4) | 0.3 (0.3-0.4) | 0.3 (0.3-0.4) |
| **LTC high 70+** | week 2 | 596 | 377 | 0.5 (0.5-0.6) | 0.5 (0.4-0.5) | 0.5 (0.4-0.5) |
| **LTC high 70+** | week 3 | 590 | 411 | 0.6 (0.5-0.6) | 0.5 (0.5-0.6) | 0.5 (0.5-0.6) |
| **LTC high 70+** | week 4 | 579 | 417 | 0.6 (0.5-0.7) | 0.6 (0.5-0.7) | 0.6 (0.5-0.7) |
| **LTC high 70+** | week 5 | 562 | 393 | 0.7 (0.6-0.8) | 0.6 (0.6-0.7) | 0.6 (0.6-0.7) |
| **LTC high 70+** | week 6 | 532 | 353 | 0.7 (0.6-0.8) | 0.6 (0.6-0.7) | 0.6 (0.6-0.7) |
| **LTC high 70+** | week 7 | 470 | 296 | 0.7 (0.6-0.8) | 0.6 (0.6-0.7) | 0.6 (0.6-0.7) |
| **LTC high 70+** | week 8 | 372 | 233 | 0.7 (0.6-0.8) | 0.6 (0.5-0.7) | 0.6 (0.5-0.7) |
| **LTC low 70+** | reference | 3485 | 1510 | [ref] | [ref] | [ref] |
| **LTC low 70+** | week 1 | 152 | 27 | 0.3 (0.2-0.4) | 0.3 (0.2-0.4) | 0.3 (0.2-0.4) |
| **LTC low 70+** | week 2 | 151 | 37 | 0.4 (0.3-0.5) | 0.4 (0.3-0.5) | 0.4 (0.3-0.5) |
| **LTC low 70+** | week 3 | 149 | 51 | 0.5 (0.4-0.7) | 0.5 (0.3-0.6) | 0.5 (0.3-0.6) |
| **LTC low 70+** | week 4 | 145 | 47 | 0.5 (0.3-0.6) | 0.4 (0.3-0.6) | 0.4 (0.3-0.6) |
| **LTC low 70+** | week 5 | 138 | 39 | 0.4 (0.3-0.6) | 0.4 (0.3-0.5) | 0.4 (0.3-0.5) |
| **LTC low 70+** | week 6 | 127 | 31 | 0.4 (0.3-0.6) | 0.4 (0.2-0.5) | 0.4 (0.2-0.5) |

| | | | | | | |
|---|---|---|---|---|---|---|
| **LTC low 70+** | week 7 | 105 | 20 | 0.3 (0.2-0.5) | 0.3 (0.2-0.5) | 0.3 (0.2-0.5) |
| **LTC low 70+** | week 8 | 73 | 24 | 0.6 (0.4-1.0) | 0.5 (0.3-0.9) | 0.5 (0.3-0.9) |
| **Disability care** | reference | 13753 | 717 | [ref] | [ref] | [ref] |
| **Disability care** | week 1 | 495 | 13 | 0.5 (0.3-0.9) | 0.4 (0.2-0.7) | 0.4 (0.2-0.7) |
| **Disability care** | week 2 | 484 | 22 | 0.9 (0.6-1.4) | 0.6 (0.4-1) | 0.7 (0.4-1.0) |
| **Disability care** | week 3 | 458 | 17 | 0.8 (0.5-1.4) | 0.5 (0.3-0.9) | 0.5 (0.3-0.9) |
| **Disability care** | week 4 | 408 | 19 | 1.1 (0.7-1.9) | 0.7 (0.4-1.1) | 0.7 (0.4-1.2) |
| **Disability care** | week 5 | 357 | 19 | 1.4 (0.8-2.4) | 0.8 (0.5-1.4) | 0.8 (0.5-1.4) |
| **Disability care** | week 6 | 295 | <10 | 0.9 (0.4-1.9) | 0.5 (0.2-1.0) | 0.5 (0.2-1.1) |
| **Disability care** | week 7 | 210 | 14 | 2.3 (1.2-4.4) | 1.1 (0.6-2.2) | 1.2 (0.7-2.3) |
| **Disability care** | week 8 | 127 | <10 | 2.1 (0.9-4.9) | 1.0 (0.4-2.2) | 1.0 (0.4-2.4) |
| **<1931 (90+)** | reference | 9566 | 3816 | [ref] | [ref] | [ref] |
| **<1931 (90+)** | week 1 | 346 | 63 | 0.2 (0.2-0.3) | 0.2 (0.2-0.3) | 0.2 (0.2-0.3) |
| **<1931 (90+)** | week 2 | 342 | 97 | 0.3 (0.2-0.4) | 0.3 (0.3-0.4) | 0.3 (0.3-0.4) |
| **<1931 (90+)** | week 3 | 336 | 89 | 0.3 (0.2-0.3) | 0.3 (0.2-0.3) | 0.3 (0.2-0.3) |
| **<1931 (90+)** | week 4 | 327 | 103 | 0.3 (0.2-0.4) | 0.3 (0.2-0.4) | 0.3 (0.2-0.4) |
| **<1931 (90+)** | week 5 | 316 | 101 | 0.3 (0.2-0.3) | 0.3 (0.2-0.3) | 0.3 (0.2-0.3) |
| **<1931 (90+)** | week 6 | 297 | 96 | 0.3 (0.2-0.3) | 0.3 (0.2-0.3) | 0.3 (0.2-0.3) |
| **<1931 (90+)** | week 7 | 267 | 85 | 0.3 (0.2-0.3) | 0.3 (0.2-0.3) | 0.3 (0.2-0.3) |
| **<1931 (90+)** | week 8 | 221 | 52 | 0.2 (0.1-0.3) | 0.2 (0.2-0.3) | 0.2 (0.2-0.3) |
| **1931-1950 (70-89)** | reference | 251070 | 20489 | [ref] | [ref] | [ref] |
| **1931-1950 (70-89)** | week 1 | 11964 | 298 | 0.2 (0.2-0.2) | 0.2 (0.2-0.2) | 0.2 (0.2-0.2) |
| **1931-1950 (70-89)** | week 2 | 11868 | 418 | 0.2 (0.2-0.3) | 0.2 (0.2-0.2) | 0.2 (0.2-0.2) |
| **1931-1950 (70-89)** | week 3 | 11665 | 495 | 0.3 (0.2-0.3) | 0.2 (0.2-0.2) | 0.2 (0.2-0.2) |
| **1931-1950 (70-89)** | week 4 | 11119 | 463 | 0.2 (0.2-0.3) | 0.2 (0.2-0.2) | 0.2 (0.2-0.2) |

| | | | | | | |
|---|---|---|---|---|---|---|
| **1931-1950 (70-89)** | week 5 | 9998 | 417 | 0.2 (0.2-0.3) | 0.2 (0.2-0.2) | 0.2 (0.2-0.2) |
| **1931-1950 (70-89)** | week 6 | 8122 | 382 | 0.3 (0.3-0.3) | 0.2 (0.2-0.2) | 0.2 (0.2-0.2) |
| **1931-1950 (70-89)** | week 7 | 5346 | 288 | 0.3 (0.3-0.4) | 0.2 (0.2-0.2) | 0.2 (0.2-0.2) |
| **1931-1950 (70-89)** | week 8 | 2533 | 159 | 0.4 (0.3-0.5) | 0.2 (0.1-0.2) | 0.2 (0.1-0.2) |
| **1951-1970 (50-69)** | reference | 363851 | 6339 | [ref] | [ref] | [ref] |
| **1951-1970 (50-69)** | week 1 | 22072 | 94 | 0.2 (0.2-0.3) | 0.2 (0.1-0.2) | 0.2 (0.2-0.2) |
| **1951-1970 (50-69)** | week 2 | 21621 | 125 | 0.3 (0.2-0.3) | 0.2 (0.2-0.3) | 0.2 (0.2-0.3) |
| **1951-1970 (50-69)** | week 3 | 20457 | 128 | 0.3 (0.2-0.3) | 0.2 (0.2-0.3) | 0.2 (0.2-0.3) |
| **1951-1970 (50-69)** | week 4 | 17578 | 118 | 0.3 (0.2-0.4) | 0.2 (0.2-0.3) | 0.2 (0.2-0.3) |
| **1951-1970 (50-69)** | week 5 | 12669 | 93 | 0.3 (0.3-0.4) | 0.2 (0.2-0.3) | 0.2 (0.2-0.3) |
| **1951-1970 (50-69)** | week 6 | 6633 | 38 | 0.3 (0.2-0.4) | 0.2 (0.1-0.2) | 0.2 (0.1-0.2) |
| **1951-1970 (50-69)** | week 7 | 2447 | 23 | 0.4 (0.3-0.6) | 0.3 (0.2-0.5) | 0.3 (0.2-0.5) |
| **1951-1970 (50-69)** | week 8 | 1104 | 18 | 0.7 (0.5-1.2) | 0.7 (0.4-1.1) | 0.7 (0.5-1.2) |
| **1971-2009 (12-49)** | reference | 383635 | 719 | [ref] | [ref] | [ref] |
| **1971-2009 (12-49)** | week 1 | 19518 | 10 | 0.3 (0.2-0.6) | 0.2 (0.1-0.4) | 0.2 (0.1-0.4) |
| **1971-2009 (12-49)** | week 2 | 18253 | 14 | 0.5 (0.3-0.9) | 0.4 (0.2-0.6) | 0.3 (0.2-0.6) |

| | | | | | | |
|---|---|---|---|---|---|---|
| **1971-2009 (12-49)** | week 3 | 15397 | <10 | 0.2 (0.1-0.5) | 0.1 (0.1-0.4) | 0.1 (0.1-0.4) |
| **1971-2009 (12-49)** | week 4 | 9390 | <10 | 0.2 (0.1-0.6) | 0.1 (0-0.4) | 0.1 (0-0.4) |
| **1971-2009 (12-49)** | week 5 | 4144 | <10 | 0.6 (0.2-1.6) | 0.4 (0.1-1.0) | 0.4 (0.1-1.0) |
| **1971-2009 (12-49)** | week 6 | 2459 | <10 | 0.2 (0-1.8) | 0.2 (0-1.4) | 0.2 (0-1.3) |
| **1971-2009 (12-49)** | week 7 | 1747 | <10 | 0.7 (0.2-2.9) | 0.6 (0.1-2.3) | 0.6 (0.1-2.2) |
| **1971-2009 (12-49)** | week 8 | 1181 | <10 | <0.01 (--) | <0.01 (--) | <0.01 (--) |
