## Supplementary file 2 for "Effect of COVID-19 vaccination on mortality by COVID-19 and on mortality by other causes, the Netherlands, January 2021- January 2022"

### Supplementary file 2 to COVID-19 vaccine effectiveness against mortality and risk of death from other causes after COVID-19 vaccination, the Netherlands, January 2021- January 2022

#### Medical risk groups

Dutch national healthcare registry data were used to define underlying medical conditions that are associated with medical risk requiring prioritization in vaccination programmes. The national registry data comprised data from an all-payer claims database of (hospital-based) outpatient specialist care utilization, and data regarding medication prescriptions at ATC-4 code level in the Netherlands. Vektis Health Care Information Center manages the all-payer claims database to inform policy decisions and for risk equalization (i.e., to prevent risk selection in the healthcare system) (Geurten, 2022; Hasaart, 2011). In the Dutch funding system of 'Diagnosis Treatment Combination' (DTC), the reimbursement per case is fixed and based on the medical cost of the bundle of healthcare services ('care activities') associated with the diagnosis classification within a set period (CBS, 2022a; Hasaart, 2011). The medication dispense database is managed by the National Health Care Institute (CBS, 2022b).

This supplementary file lists the specific conditions that fall under two types of medical risk groups (i.e., high and intermediate medical risk), and lists the definitions that were made within the registry datasets to approximate these conditions. The medical high-risk group was defined based on conditions associated with high risk for severe COVID-19. These patients were prioritized for primary series vaccination in 2021. The medical intermediate-risk group was based on the eligibility criteria for influenza vaccination in the Netherlands. Both groups are described in the COVID-19 vaccination guideline of the Dutch National Public Health Institute (RIVM, 2022). For both groups, age and long-term care use are part of the criteria, but these variables were excluded from the definition of medical risk in the current study, as they were already included as time-varying variables.

Within the two data sources either one, or, a combination of three variables (DTC codes, care activity codes, and dispensed medications) were used to identify individuals within the medical risk groups. The individual-level, pseudonymized data were available within the secured working environment of Statistics Netherlands.

1) Outpatient specialist care utilization – Vektis

   In the DTC data, we used the codes of the DTC product (7- or 8-digit codes) and care activities (6-digit codes) that were registered by medical specialists for the purpose of health insurance claims. The DTC product code relates to a (diagnosis) classification within a specific medical specialty (i.e., classifications are not uniform between medical specialties) and range from broad to narrow definitions. A care activity is more narrow-defined and is not (necessarily) specific to one type of DTC product.

   With these DTC and care activity codes, we were able to identify individuals that visited a medical specialist in a given year for the treatment of relevant medical conditions. A descriptive list of DTC products and care activities is available, in Dutch, within the CBS (Statistics Netherlands) open data environment that can be accessed through the following URL: Indeling van DBC-diagnosen in de MSZ, 2019-2021 (cbs.nl)

2) Medication dispensing data – National Health Care Institute

   Data on medication included all medication dispensed and covered by the statutory basic health insurance in the Netherlands. Medication received in hospitals and nursing homes are not included in the database. Dispensed medication is classified into ATC-4 codes according to the WHO drug classification system. The ATC-classification can be found at: WHOCC - Home

Table S2.1 Overview of medical risk groups based on national healthcare registry data

| | Years of data included | DTC product codes | | Care activity codes | Medication (ATC-4 code) |
|---|---|---|---|---|---|
| | | *Specialism* | *Diagnosis* | | |
| **Medical high-risk group** | | | | | |
| Hematologic malignancies | 2016-2020 | 313 | 0751 | | |
| | | 313 | 0752 | | |
| | | 313 | 0753 | | |
| | | 313 | 0754 | | |
| | | 313 | 0756 | | |
| | | 313 | 0757 | | |
| | | 313 | 0761 | | |
| | | 313 | 0771 | | |
| | | 313 | 0773 | | |
| | | 316 | 6101 | | |
| | | 316 | 6102 | | |
| | | 316 | 6106 | | |
| | | 316 | 6111 | | |
| | | 361 | 0110 | | |
| Sickle cell disease | 2016-2020 | 313 | 0703 | | |
| Severe renal failure (with dialysis)[a] | 2020 | 303 | 0435 | 033686 | |
| | | 313 | 0322 | 033687 | |
| | | 313 | 0326 | 033702 | |
| | | 313 | 0331 | 033703 | |
| | | 313 | 0332 | 080832 | |
| | | 313 | 0333 | 190156 | |
| | | 313 | 0334 | 192048 | |
| | | 313 | 0335 | 192049 | |
| | | 313 | 0336 | 192051 | |
| | | 313 | 0337 | 192052 | |
| | | 313 | 0338 | 192053 | |
| | | 313 | 0339 | 192054 | |
| | | 328 | 2270 | 192055 | |
| | | 362 | 0225 | 192056 | |
| | | 362 | 0226 | 192058 | |
| | | | | 192059 | |
| | | | | 192061 | |
| | | | | 192062 | |
| | | | | 192063 | |
| | | | | 192064 | |
| | | | | 192065 | |
| | | | | 192066 | |
| | | | | 192067 | |
| | | | | 192068 | |
| | | | | 192069 | |

| | | | |
|---|---|---|---|
| | | | 192070 |
| Organ, stem cell, or bone marrow transplantation | 2020 | 303 0501 | 032707 |
| | | 303 0512 | 032708 |
| | | 303 0521 | 032709 |
| | | 303 0522 | 032710 |
| | | 303 0531 | 039192 |
| | | 303 0541 | 039214 |
| | | 303 0551 | 039215 |
| | | 303 0553 | 039216 |
| | | 303 0554 | 039233 |
| | | 303 0555 | 039234 |
| | | 303 0557 | 039235 |
| | | 303 0559 | 039236 |
| | | 303 0560 | 039237 |
| | | 303 0561 | 039284 |
| | | 303 0562 | 039350 |
| | | 303 0563 | 039351 |
| | | 303 0564 | 039394 |
| | | 313 0070 | 039395 |
| | | 313 0072 | 039396 |
| | | 313 0073 | 039981 |
| | | 313 0074 | 192002 |
| | | 313 0076 | 192025 |
| | | 313 0078 | 192028 |
| | | 313 0079 | 192043 |
| | | 313 0081 | 192045 |
| | | 313 0082 | 192047 |
| | | 313 0083 | 192079 |
| | | 313 0084 | 192080 |
| | | 313 0341 | 192086 |
| | | 313 0342 | 192087 |
| | | 313 0344 | 192089 |
| | | 313 0345 | 192094 |
| | | 313 0346 | 192095 |
| | | 313 0347 | 192096 |
| | | 316 7901 | 192097 |
| | | 316 7903 | 192098 |
| | | 316 7904 | 192099 |
| | | 316 7905 | 192107 |
| | | 316 7907 | 192108 |
| | | 316 7909 | 192109 |
| | | 316 7910 | 192112 |
| | | 316 7911 | 192113 |
| | | 316 7920 | 192114 |
| | | 316 7921 | 192117 |
| | | 316 7922 | 192119 |
| | | 316 7923 | 192126 |
| | | 316 7924 | |

| | | | |
|---|---|---|---|
| | | 316 7926<br>318 0715<br>318 0716<br>318 0717<br>318 0761<br>318 0763<br>318 0764<br>318 0766<br>318 0767<br>318 0768<br>318 0769<br>320 0805<br>320 0903<br>320 0904<br>320 0912<br>322 2201<br>322 2202<br>328 2910<br>328 2920<br>328 2930<br>362 0400 | |
| Severe congenital immune disorder (primary immune deficiency) | 2016-2020 | 313 0493<br>313 0711<br>316 6201<br>316 6202<br>316 6299<br>316 6203<br>326 00H4<br>326 00H5 | |
| Neurological disorders with compromised breathing | 2020 | 322 2102<br>322 2104<br>330 0522<br>330 0901<br>330 0911 | |
| A solid tumor (with chemotherapy and/or radiotherapy) | 2020 | | 032701<br>032717<br>034730<br>039141<br>039142<br>039143<br>039145<br>039886<br>039888<br>039891<br>039892<br>039893<br>039895<br>039958 |

| | | | | |
|---|---|---|---|---|
| | | | 090791 | |
| | | | 090792 | |
| | | | 090796 | |
| | | | 090797 | |
| | | | 090830 | |
| | | | 090833 | |
| Immunosuppressants | 2020 | | | L04A |
| Morbid obesity | 2020 | 303 0342 | | |
| | | 303 0341 | | |
| | | 313 0283 | | |
| | | 316 7101 | | |
| **Medical intermediate-risk group** | | | | |
| Lung disease | 2020 | 302 0031* | | RO3A |
| | | 303 0178* | | |
| | | 303 0309 | | |
| | | 303 0312* | | |
| | | 303 0313 | | |
| | | 313 0007* | | |
| | | 313 0125* | | |
| | | 313 0401* | | |
| | | 313 0402* | | |
| | | 313 0409* | | |
| | | 313 0601 | | |
| | | 313 0609* | | |
| | | 316 3103* | | |
| | | 316 3109* | | |
| | | 316 3202 | | |
| | | 316 3203* | | |
| | | 316 3207* | | |
| | | 316 3208* | | |
| | | 316 3299* | | |
| | | 316 7706 | | |
| | | 322 1101* | | |
| | | 322 1103* | | |
| | | 322 1104* | | |
| | | 322 1201 | | |
| | | 322 1202 | | |
| | | 322 1241 | | |
| | | 322 1303 | | |
| | | 322 1304 | | |
| | | 322 1305 | | |
| | | 322 1307 | | |
| | | 322 1308 | | |
| | | 322 1401* | | |
| | | 322 1403 | | |
| | | 322 1404 | | |
| | | 322 1405* | | |
| | | 322 1511 | | |

| | | | | |
|---|---|---|---|---|
| | | 322 1513 | | |
| | | 322 1514 | | |
| | | 322 1601* | | |
| | | 322 1602 | | |
| | | 322 1603* | | |
| | | 322 1801* | | |
| | | 322 1803* | | |
| | | 322 2102 | | |
| | | 322 2104 | | |
| | | 322 9906* | | |
| | | 326 00B1* | | |
| | | 326 00B2* | | |
| | | 326 00B3* | | |
| | | 326 00B4* | | |
| | | 326 00C3* | | |
| | | 326 00C4* | | |
| | | 326 00G1* | | |
| | | 326 00G3* | | |
| | | 327 0615* | | |
| | | 328 1260* | | |
| | | 328 1320* | | |
| | | 328 1330* | | |
| | | 328 1350* | | |
| | | 328 1450* | | |
| | | 328 1480 | | |
| | | 328 1570* | | |
| | | 328 1590 | | |
| | | 328 1595 | | |
| | | 335 0271* | | |
| | | 335 0272 | | |
| | | 335 0273* | | |
| | | 8418 0503* | | |
| Chronic cardiac disease | 2020 | 303 0178* | | C01A |
| | | 313 0119* | | C01B |
| | | 313 0125* | | C01C |
| | | 313 0101 | | C01D |
| | | 313 0102 | | C01E |
| | | 313 0106 | | |
| | | 313 0107 | | |
| | | 316 3408* | | |
| | | 316 3411* | | |
| | | 316 3499* | | |
| | | 316 3406 | | |
| | | 316 3410 | | |
| | | 316 7920 | | |
| | | 316 7922 | | |
| | | 318 0907* | | |
| | | 320 0301* | | |

| | | |
|---|---|---|
| | | 320 0810* |
| | | 320 0822* |
| | | 320 0911* |
| | | 320 0202 |
| | | 320 0203 |
| | | 320 0204 |
| | | 320 0205 |
| | | 320 0302 |
| | | 320 0401 |
| | | 320 0402 |
| | | 320 0403 |
| | | 320 0404 |
| | | 320 0409 |
| | | 320 0501 |
| | | 320 0509 |
| | | 320 0801 |
| | | 320 0802 |
| | | 320 0803 |
| | | 320 0804 |
| | | 320 0805 |
| | | 320 0806 |
| | | 320 0821 |
| | | 320 0903 |
| | | 320 0904 |
| | | 320 0912 |
| | | 322 1803* |
| | | 322 2101 |
| | | 322 2202 |
| | | 327 0613* |
| | | 328 2110* |
| | | 328 2120* |
| | | 328 2130* |
| | | 328 2220 |
| | | 328 2230 |
| | | 328 2240 |
| | | 328 2260 |
| | | 328 2320 |
| | | 328 2325 |
| | | 328 2335 |
| | | 328 2340 |
| | | 328 2360 |
| | | 328 2390 |
| | | 328 2400 |
| | | 328 2405 |
| | | 328 2410 |
| | | 328 2415 |
| | | 328 2420 |
| | | 328 2425 |

| | | | |
|---|---|---|---|
| | | 328 | 2430 |
| | | 328 | 2450 |
| | | 328 | 2455 |
| | | 328 | 2460 |
| | | 328 | 2465 |
| | | 328 | 2470 |
| | | 328 | 2475 |
| | | 328 | 2510 |
| | | 328 | 2520 |
| | | 328 | 2525 |
| | | 328 | 2530 |
| | | 328 | 2543 |
| | | 328 | 2545 |
| | | 328 | 2550 |
| | | 328 | 2555 |
| | | 328 | 2560 |
| | | 328 | 2565 |
| | | 328 | 2570 |
| | | 328 | 2575 |
| | | 328 | 2580 |
| | | 328 | 2585 |
| | | 328 | 2590 |
| | | 328 | 2610 |
| | | 328 | 2630 |
| | | 328 | 2635 |
| | | 328 | 2640 |
| | | 328 | 2645 |
| | | 328 | 2650 |
| | | 328 | 2655 |
| | | 328 | 2660 |
| | | 328 | 2665 |
| | | 328 | 2675 |
| | | 328 | 2680 |
| | | 328 | 2695 |
| | | 328 | 2720 |
| | | 328 | 2730 |
| | | 328 | 2740 |
| | | 328 | 2750 |
| | | 328 | 2760 |
| | | 328 | 2770 |
| | | 328 | 2785 |
| | | 328 | 2790 |
| | | 328 | 2910 |
| | | 328 | 2930 |
| | | 328 | 2940 |
| | | 335 | 0261* |
| | | 335 | 0262 |
| | | 362 | 0311* |

| | | | | |
|---|---|---|---|---|
| | | 8418 0501* | | |
| Diabetes Mellitus | 2020 | 301 0754 | 035520 | A10A |
| | | 301 0755 | 035523 | A10B |
| | | 301 0757 | 035524 | A10X |
| | | 301 0759 | 035525 | |
| | | 303 0432 | 039582 | |
| | | 303 0522 | 039583 | |
| | | 303 0553 | 039641 | |
| | | 305 2065 | 190309 | |
| | | 313 0072 | 190351 | |
| | | 313 0221 | | |
| | | 313 0222 | | |
| | | 313 0223 | | |
| | | 316 7104 | | |
| | | 316 7113 | | |
| | | 316 7114 | | |
| | | 316 7903 | | |
| | | 318 0902 | | |
| | | 335 0222 | | |
| Chronic renal failure | 2020 | 303 0435 | 033686 | |
| | | 313 0304 | 033687 | |
| | | 313 0313 | 033702 | |
| | | 313 0324 | 033703 | |
| | | 313 0325 | 080832 | |
| | | 313 0326 | 190156 | |
| | | 313 0331 | 192048 | |
| | | 313 0332 | 192049 | |
| | | 313 0333 | 192051 | |
| | | 313 0334 | 192052 | |
| | | 313 0335 | 192053 | |
| | | 313 0336 | 192054 | |
| | | 313 0337 | 192055 | |
| | | 313 0338 | 192056 | |
| | | 313 0339 | 192058 | |
| | | 313 0342 | 192059 | |
| | | 316 4004 | 192061 | |
| | | 316 4006 | 192062 | |
| | | 328 2270 | 192063 | |
| | | 362 0225 | 192064 | |
| | | 362 0226 | 192065 | |
| | | | 192066 | |
| | | | 192067 | |
| | | | 192068 | |
| | | | 192069 | |
| | | | 192070 | |
| Bone marrow transplantation | 2020 | | 032707 | |
| | | | 032708 | |
| | | | 032709 | |

| | | | | |
|---|---|---|---|---|
| | | | 032710<br>039284<br>039981<br>192079<br>192080<br>192086<br>192087<br>192089<br>192094<br>192095<br>192096<br>192097<br>192098<br>192099<br>192117<br>192119 | |
| Human immunodeficiency virus (hiv) | 2016-2020 | 313 0461<br>313 0462<br>316 7802<br>322 1513<br>322 1522 | | |
| Reduced immunity against infection | 2020 As | 303 0325*<br>303 0326*<br>303 0501<br>303 0512<br>303 0521<br>303 0522<br>303 0531<br>303 0541<br>303 0551<br>303 0553<br>303 0554<br>303 0555<br>303 0557<br>303 0559<br>303 0560<br>303 0561<br>303 0562<br>303 0563<br>303 0564<br>303 0836*<br>304 0386*<br>305 1052*<br>305 1680*<br>305 2070*<br>313 0070<br>313 0072<br>313 0073 | 032701<br>032717<br>034730<br>039141<br>039142<br>039143<br>039145<br>039886<br>039888<br>039891<br>039892<br>039893<br>039895<br>039958<br>090791<br>090792<br>090796<br>090797<br>090830<br>090833<br>032707<br>032708<br>032709<br>032710<br>039192<br>039214<br>039215 | L04A<br>A07E^<br>D05B^<br>H02A^<br>H02B^<br>M01A^<br>M01C^<br>M02A^<br>M04A^ |

| | | | |
|---|---|---|---|
| | | 313 0074 | 039216 |
| | | 313 0076 | 039233 |
| | | 313 0078 | 039234 |
| | | 313 0079 | 039235 |
| | | 313 0081 | 039236 |
| | | 313 0082 | 039237 |
| | | 313 0083 | 039284 |
| | | 313 0084 | 039350 |
| | | 313 0341 | 039351 |
| | | 313 0342 | 039394 |
| | | 313 0344 | 039395 |
| | | 313 0345 | 039396 |
| | | 313 0346 | 039981 |
| | | 313 0347 | 192002 |
| | | 313 0461 | 192025 |
| | | 313 0462 | 192028 |
| | | 313 0493 | 192043 |
| | | 313 0503 | 192045 |
| | | 313 0505* | 192047 |
| | | 313 0506* | 192079 |
| | | 313 0507 | 192080 |
| | | 313 0512* | 192086 |
| | | 313 0521* | 192087 |
| | | 313 0522 | 192089 |
| | | 313 0525 | 192094 |
| | | 313 0526* | 192095 |
| | | 313 0527* | 192096 |
| | | 313 0531 | 192097 |
| | | 313 0751 | 192098 |
| | | 313 0752 | 192099 |
| | | 313 0753 | 192107 |
| | | 313 0754 | 192108 |
| | | 313 0756 | 192109 |
| | | 313 0757 | 192112 |
| | | 313 0761 | 192113 |
| | | 313 0763 | 192114 |
| | | 313 0771 | 192117 |
| | | 313 0773 | 192119 |
| | | 313 0774 | 192126 |
| | | 313 0922* | |
| | | 313 0923* | |
| | | 313 0941 | |
| | | 313 0943 | |
| | | 313 0945 | |
| | | 313 0946 | |
| | | 316 3205 | |
| | | 316 3314* | |
| | | 316 5001* | |

| | | |
|---|---|---|
| | | 316 5003* |
| | | 316 5004 |
| | | 316 5099* |
| | | 316 6101 |
| | | 316 6102 |
| | | 316 6104 |
| | | 316 6106 |
| | | 316 6111 |
| | | 316 6112 |
| | | 316 6124 |
| | | 316 6201 |
| | | 316 6202 |
| | | 316 6203 |
| | | 316 7501 |
| | | 316 7802 |
| | | 316 7901 |
| | | 316 7903 |
| | | 316 7904 |
| | | 316 7905 |
| | | 316 7907 |
| | | 316 7909 |
| | | 316 7910 |
| | | 316 7911 |
| | | 316 7920 |
| | | 316 7921 |
| | | 316 7922 |
| | | 316 7923 |
| | | 316 7924 |
| | | 316 7926 |
| | | 318 0408 |
| | | 318 0601 |
| | | 318 0602* |
| | | 318 0707 |
| | | 318 0708 |
| | | 318 0709 |
| | | 318 0713 |
| | | 318 0715 |
| | | 318 0716 |
| | | 318 0717 |
| | | 318 0718 |
| | | 318 0761 |
| | | 318 0763 |
| | | 318 0764 |
| | | 318 0766 |
| | | 318 0767 |
| | | 318 0768 |
| | | 318 0769 |
| | | 320 0805 |

| | | |
|---|---|---|
| | | 320 0903 |
| | | 320 0904 |
| | | 320 0912 |
| | | 322 1403 |
| | | 322 1513 |
| | | 322 1522 |
| | | 322 2201 |
| | | 322 2202 |
| | | 324 0101* |
| | | 324 0102* |
| | | 324 0107* |
| | | 324 0112* |
| | | 324 0202* |
| | | 324 0301 |
| | | 324 0302 |
| | | 324 0303 |
| | | 324 0304 |
| | | 324 0305 |
| | | 324 0309* |
| | | 324 0310* |
| | | 324 0311* |
| | | 324 0312 |
| | | 324 0313 |
| | | 324 0315 |
| | | 324 0317 |
| | | 324 0319 |
| | | 324 0399* |
| | | 326 00H4 |
| | | 327 0117* |
| | | 328 2910 |
| | | 328 2920 |
| | | 328 2930 |
| | | 361 0110 |
| | | 362 0400 |
| | | 8418 0508* |
| Neuro-muscular disease | 2020 | 303 0402 |
| | | 305 1054 |
| | | 308 1201 |
| | | 308 1205 |
| | | 308 1210 |
| | | 308 1220 |
| | | 308 1225 |
| | | 308 1230 |
| | | 308 1235 |
| | | 308 1240 |
| | | 308 1321 |
| | | 308 1515 |
| | | 316 3501 |

| | | | |
|---|---|---|---|
| | | 316 3503<br>316 3508<br>316 7708<br>322 2102<br>322 2104<br>327 0313<br>328 3210<br>328 3310<br>330 0501<br>330 0503<br>330 0511<br>330 0522<br>330 0531<br>330 0601<br>330 0602<br>330 0621<br>330 0901<br>330 0911<br>330 0999<br>330 1101<br>330 1102<br>330 1103<br>330 1111<br>330 1121<br>330 1199<br>330 9901<br>330 9920<br>330 9925<br>330 9926<br>335 0252<br>362 0101<br>362 0102<br>362 0103<br>362 0111<br>362 0112<br>362 0113<br>362 0114<br>362 0115<br>362 0201<br>362 0202<br>362 0203<br>8418 0101 | |
| Cochlear implants | 2016-2020 | | 031907<br>031908 |

[a] For severe renal failure, DTC products were only included if at least one care activity in the same category was included. Care activities were only included if at least one DTC product in the same category was included
* DTC product only included when ATC-4 medication code in the same category was also included

^ Medication code was only included when DTC product in the same category was also included

**References**

Geurten, R. J., Struijs, J. N., Elissen, A. M., Bilo, H. J., van Tilburg, C., & Ruwaard, D. (2022). Delineating the Type 2 Diabetes Population in the Netherlands Using an All-Payer Claims Database: Specialist Care, Medication Utilization and Expenditures 2016–2018. *PharmacoEconomics-open*, *6*(2), 219-229.

Hasaart, F. (2011). *Incentives in the diagnosis treatment combination payment system for specialist medical care : a study about behavioral responses of medical specialists and hospitals in the Netherlands.* [Doctoral Thesis, Maastricht University]. Maastricht University. https://doi.org/10.26481/dis.20111104fh

RIVM, National Institute for Public Health and the Environment (2022a). COVID-19-vaccinatie uitvoeringsrichtlijn. [COVID-19 vaccination implementing guideline] https://lci.rivm.nl/richtlijnen/covid-19-vaccinatie

Statistics Netherlands (CBS) (2022A). *Diagnosis and treatment combinations*. https://www.cbs.nl/en-gb/news/2010/45/diagnosis-and-treatment-combinations

Statistics Netherlands (CBS) (2022B). *Persons with dispensed medicines.* Statistics Netherlands (CBS) (2022a). *Diagnosis and treatment combinations.* https://www.cbs.nl/en-gb/news/2010/45/diagnosis-and-treatment-combinations

Statistics Netherlands (CBS) (2022b). *Persons with dispensed medicines.* https://www.cbs.nl/en-gb/our-services/methods/surveys/brief-survey-description/persons-with-dispensed-medicines
